# Frequency dynamics predict viral fitness, antigenic relationships and epidemic growth

**DOI:** 10.1101/2024.12.02.24318334

**Authors:** Marlin D. Figgins, Trevor Bedford

## Abstract

During the COVID-19 pandemic, SARS-CoV-2 variants drove large waves of infections, fueled by increased transmissibility and immune escape. Current models focus on changes in variant frequencies without linking them to underlying transmission mechanisms of intrinsic transmissibility and immune escape. We introduce a framework connecting variant dynamics to these mechanisms, showing how host population immunity interacts with viral transmissibility and immune escape to determine relative variant fitness. We advance a selective pressure metric that provides an early signal of epidemic growth using genetic data alone, crucial with current underreporting of cases. Additionally, we show that a latent immunity space model approximates immunological distances, offering insights into population susceptibility and immune evasion. These insights refine real-time forecasting and lay the groundwork for research into the interplay between viral genetics, immunity, and epidemic growth.

## Introduction

The COVID-19 pandemic was marked by the successive emergence of SARS-CoV-2 variant viruses, driving repeated epidemics globally [1, 2]. While these repeated large waves occurred with the emergence of novel variants, the mechanism driving these variants’ success changed over time. The spread of early variants such as Alpha, Beta, Gamma and Delta were largely driven by increases in intrinsic transmissibility [3]. The Omicron variant showed substantial immune escape [3] and subsequent derived lineages within Omicron including XBB, EG.5.1 and JN.1 appear to be driven by immune escape as evidenced through molecular studies of neutralization using human sera [4–7]. Since 2022, there has been repeated replacement by subsequent Omicron-derived lineages. This rapid viral population turnover is consistent with antigenic evolution and is observed in other viruses such as seasonal influenza [8], although SARS-CoV-2 currently remains an outlier in terms of pace of its evolution [9]. This transition from transmissibility-driven to immune escape-driven success is a consequence of the interplay between population immunity and variant fitness.

With the increased temporal and geographical scale of sequencing alongside a detailed genetic nomenclature [10] and bioinformatic tools for lineage assignment [11, 12], we have gained more data for SARS-CoV-2 than for other circulating viruses giving a unique opportunity for insight into its evolution. Several models of variant frequency have been developed to estimate the fitness of emerging SARS-CoV-2 variants [13–18]. These models estimate the relative fitness (or selective advantage) of circulating variant viruses from their frequency in sequencing data, typically represented by counts of variant sequences over time within a geographic region. Relative fitness in these models is often assumed to be constant and intrinsic to the variant of interest. However, this may be an oversimplification of the transmission process.

It has been shown that these transmission advantages differ geographically and temporally, suggesting that variant transmission advantages are not necessarily fixed and may be informed by regional population differences [15, 19]. In fact, heterogeneity in transmission advantages may be well explained by regional differences in immune structure as Dadonaite et al. [20] show deep mutational scanning estimates of immune escape are well correlated with estimated variant growth advantages. Existing models that allow variant transmission advantages to change in time often do not have a mechanistic underpinning for why transmission advantages exist and vary geographically and temporally [15,16]. Models that do include mechanistic grounding of transmission based on population immunity such as Meijers et al. [21] and Raharinirina et al. [22] have parameterized variant-specific immunity based on serological measurements or deep mutational scanning datasets. Timely experimental data is thus a requirement for these models to perform well for real-time evolutionary forecasting.

In response to this gap, we introduce a novel framework that links variant dynamics directly to transmission mechanisms using compartmental models of infectious diseases (Fig. 1). By modeling both intrinsic transmissibility and immune escape, we explain how shifts in population immunity shape the relative fitness of viral variants and select for immune escape over intrinsic transmissibility with increasing past exposure. Furthermore, including these mechanisms suggests that relative fitness varies in time, reflecting the evolving landscape of population immunity and exposure regardless of the underlying mechanism.

**Fig. 1.**
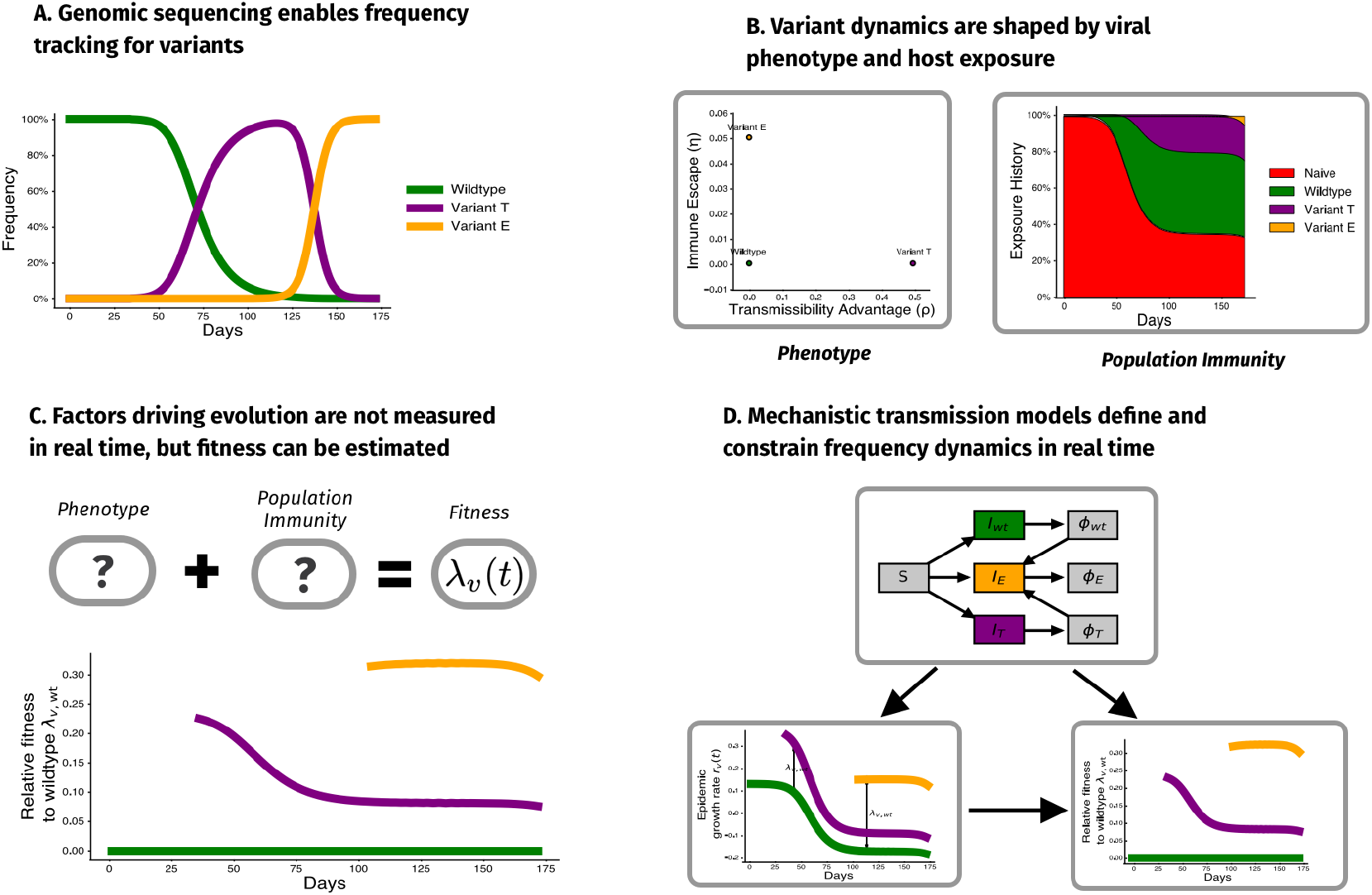
Mechanistic transmission models inform frequency dynamics. (A) Genomic surveillance reveals changes in the frequency of genetic variations over time. (B) These frequency changes can arise due to differences in phenotypes related to transmission (e.g, immune escape, transmissibility, binding) and changes in population immunity due to recent exposure. (C) Despite being instrumental for real-time analysis, both variant phenotype and population immunity are rarely observed in real time. (D) We use mechanistic transmission models to infer relative fitness from frequency data alone, taking advantage of known structure in transmission dynamics. This enables us to quantify trade-offs between variant phenotypes and develop new methods for estimating fitness in populations undergoing antigenic evolution.

Here, we present a novel non-parametric method for estimating time-varying fitness regardless of the underlying transmission mechanism. Alongside this development, we introduce a “selective pressure” metric that quantifies the impact of variant turnover on population-level epidemic growth rates. Finally, we develop a latent immunity model that we use to estimate the underlying proportion of pseudo-immune groups within multiple geographies and pseudo-immune escape rates for circulating variants that predicts antigenic distances using sequence data alone. Overall, our framework bridges the gap between genetic data and transmission dynamics, offering a new way to predict and manage viral outbreaks.

## Results

### Variant dynamics and relative fitness in multistrain models

Multi-strain models of epidemics have been developed to understand the competition between different viral strains that exhibit different levels of cross-immunity [23, 24]. These models have typically been used to explain strain evolution in antigenically variable pathogens like seasonal influenza virus [8] and seasonal coronaviruses [25, 26].

We begin by modeling a population of *V* exponentially growing variant viruses *v* each with prevalence *I*_*v*_(*t*) and time-varying growth rate *r*_*v*_(*t*). By considering the difference in growth rates for variants *v* and *u*, we can define the relative fitness as *λ*_*v,u*_(*t*) = *r*_*v*_(*t*) − *r*_*u*_(*t*). This relative fitness determines the change in the frequencies of the variants in the population

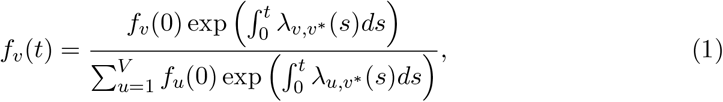

where *v*^∗^ is a chosen pivot variant that has relative fitness zero.

In order to better understand frequency dynamics of pathogens with multiple co-circulating variants, we apply the above framework to compartmental models of epidemics, which can be written as time-varying exponential growth (detailed in Supplementary Text S1). These models provide an intuition of how strain-level selection depends on the assumed transmission mechanism of the underlying epidemic model. This framework also generalizes several existing methods for relative fitness estimation and prediction (detailed in Supplementary Text S2). We summarize dynamics of a three-variant mechanistic transmission model in Fig. S1, where we compare a transmission variant *T* with a 50% increase in transmissibility (*ρ* = 0.5) to an escape variant *E* that infects 5% of hosts possessing wildtype immunity (*η* = 0.05).

### Determining the transmissibility-escape tradeoff

To understand the fitness trade-off between transmissibility and immune escape, we consider dynamics with a wildtype virus *W* with *ρ*_*W*_ = 0 and *η*_*W*_ = 0, an increased transmissibility variant *T* with *ρ* = *ρ*_*T*_ > 0 and *η* = *η*_*T*_ = 0 and an immune escape variant *E* with *ρ*_*E*_ = 0 and *η*_*E*_ > 0.

Following Equation 35, we write relative fitnesses of the escape variant or transmissibility variant as

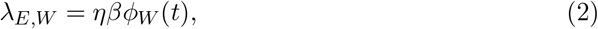

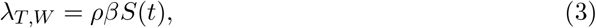

where *β* is the transmissibility coefficient, *η* is escape proportion against the wildtype, *ρ* is the variant’s proportional increase to transmissibility, and *S*(*t*) and *ϕ*_*W*_ are the proportions of the population who are susceptible and have wildtype immunity respectively.

In the simplest case where individuals are either susceptible or have wildtype immunity (*S*(*t*) + *ϕ*_*W*_ (*t*) = 1), we can compute the critical immune fraction *ϕ*^∗^ at which *λ*_*E,W*_ (*ϕ*^∗^) = *λ*_*T,W*_ (*ϕ*^∗^) as

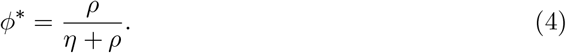

For past exposure level greater than *ϕ*^∗^ escape variants have a higher relative fitness. This trade off shows that increasing degree of escape entails that a lower proportion of past exposure is needed for escape variants to be preferred (Fig. 2). Additionally, this shows that when intrinsic transmissibility increases are limited escape is more likely to be a dominant mechanism for variant turnover.

**Fig. 2.**
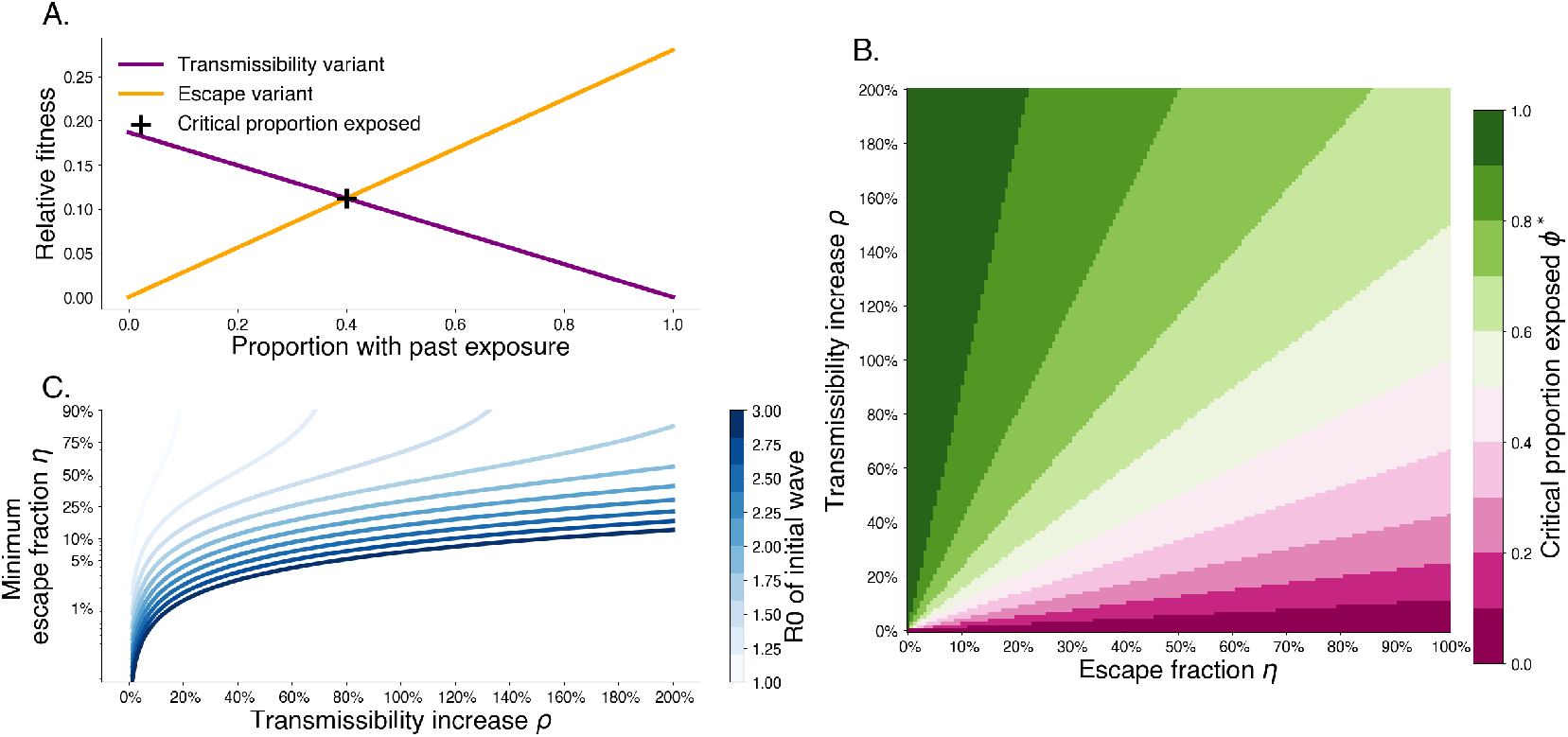
Trade-off between degree of immune escape and increased transmissibility. A. Relative fitness for a transmissibility increasing variant *T* with *ρ* = 0.2 and an immune escaping variant *E* with *η* = 0.3 for *R*_0,*W*_ = 2.8 and 1*/γ* = 3.0 days. The intersection point shows that after 40% of the population has wildtype immunity, the escape variant has higher fitness. B. The critical exposure proportion is shown for various escape fraction and transmissibility increase. Above the critical exposure proportion, we expect dominance of escape variants. C. The minimum escape fraction needed for second waves to be comprised of escape variant assuming competition with transmissibility increase variants and first wave with a given *R*_0_.

### Initial growth rates insufficient for predicting short-term frequency growth

One question of interest is whether knowledge of mechanism meaningfully informs our ability to forecast short-term frequency growth. The first step to addressing this is to understand how the relative fitness may change in time to understand the predictability of relative fitness in the short-term.

We find that the mechanistic forms analyzed in this paper (Supplementary Text S1) can be represented as weighted combinations of *B* time-varying functions ϒ_*b*_(*t*) with weights 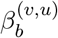. We can think of each of these functions ϒ_*b*_ as an immune background and the coefficient 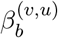 as a transmission differential between variants *v* and *u*, so that

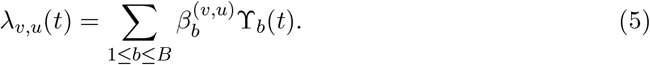

Even in the case of complete knowledge of the relative fitness and the underlying fitness contributions in the present and past, we have that change in the relative fitness is determined by

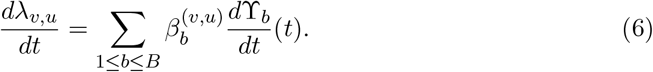

By considering a Taylor expansion of the relative fitness about the point of estimation *t*_0_, we can approximate the relative fitness in the future as

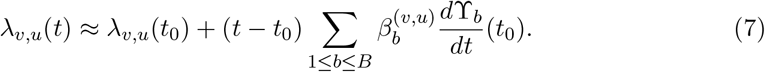

This suggests small differences in the form of *λ*_*v,u*_(*t*) can lead to meaningful differences in the future relative fitnesses through changes in the underlying immune backgrounds.

We investigate whether relative fitnesses vary predictably in the short-term regardless of mechanism. To do so, we apply the two-variant model developed in previous sections for different mechanisms of immune escape and increased transmissibility. We fix the relative fitness of the novel variant at a prediction time *t*_0_ using Equation 4 and assess the change in the relative fitness in the short-term. We find that although relative fitness trajectories share the same decreasing shape, they may decline at different rates depending on the mechanism (Fig. S3). This can lead to substantial changes in the predicted incidence depending on the assumed mechanism and affects to overall rate of turnover.

### Correlations are insufficient for mechanism identification

Although correlations between vaccination uptake and variant growth advantage are often observed, these alone may not be sufficient to identify the mechanism behind a variant’s success. A variant’s fitness advantage may arise from increased transmissibility, immune escape, or a combination of both. Even in the absence of immune escape, the relative fitness of a variant depends on the proportion of the population that is susceptible to infection and therefore changes with both past exposure and vaccine uptake (Supplementary Text S1). To illustrate this, we simulate the spread of a variant with increased transmissibility in populations with varying initial vaccination levels.

In populations with lower vaccination levels, the variant’s prevalence peaks more sharply and its relative fitness declines quickly as immunity accumulates within the population (Fig. 3A-C). In contrast, higher vaccination levels constrain relative fitness, leading to a delayed peak in prevalence and more stable relative fitness as the existing immunity limits the variant’s spread (Fig. 3A-C). Even without immune escape, estimated growth advantages for this variant decrease with increasing vaccination uptake near the beginning of an epidemic (Fig. 3D). Later in the epidemic, this relationship reverses with estimated growth advantages over the full period increasing with initial vaccination levels, which may be mistaken as signal for immune escape (Fig. 3E).

**Fig. 3.**
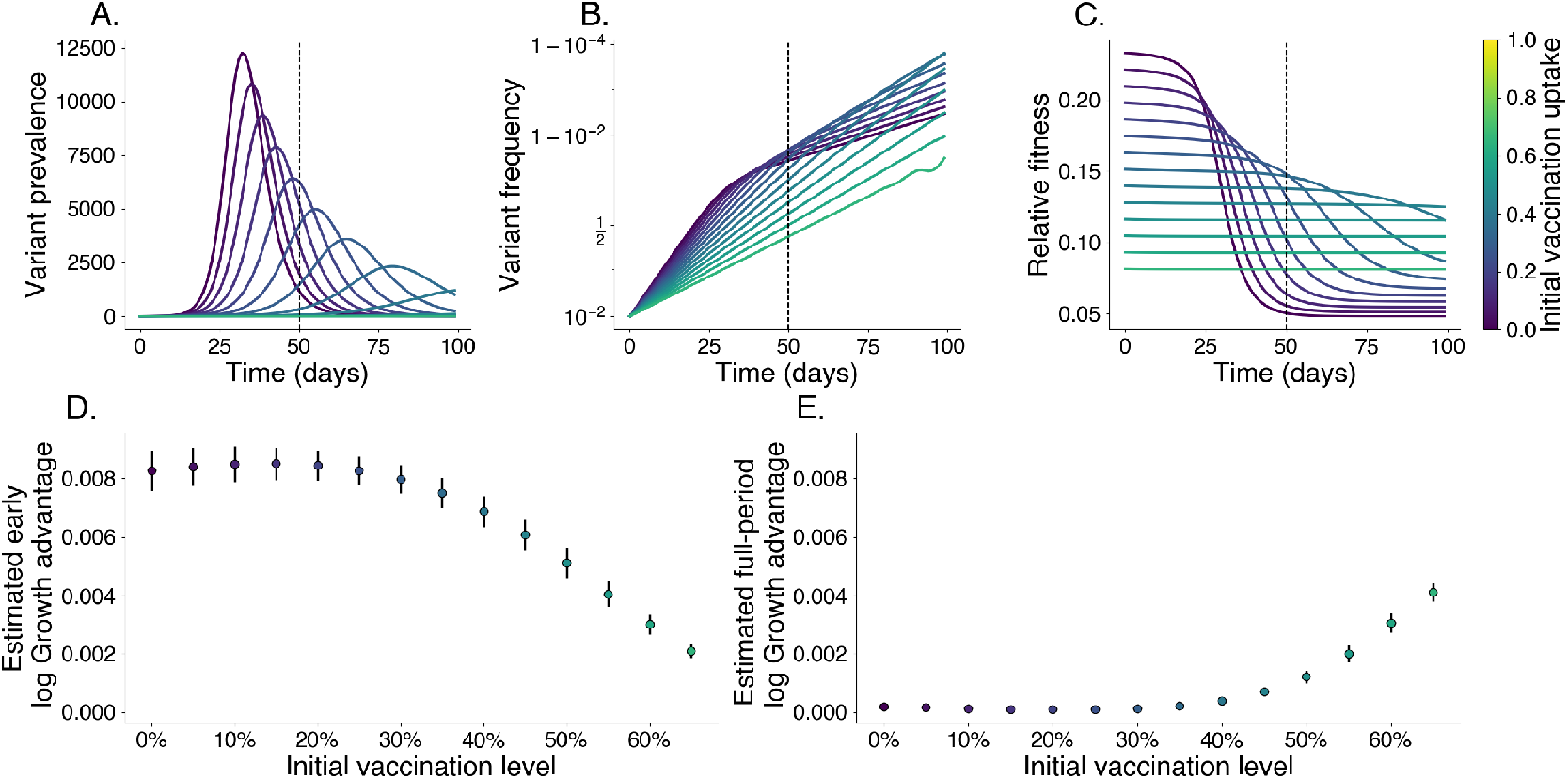
Relative fitness is correlated with vaccination levels in the absence of immune escape. We simulate the growth of a pure transmissibility increased variant at varying levels of vaccination. Darker colors represent lower vaccine uptake. We identify an early growth period where relative fitness is at its highest; the cutoff for this period is denoted with a vertical dashed line. A. Prevalence of variant, each line is its own simulation. B. Frequency of variant. C. Relative fitness for variant over time. D. Estimated log growth advantage using linear regression of log relative frequency of variant over wildtype using only data before the early cutoff. E. Same as D. but using data from the entire period shown.

This analysis shows that correlation-based methods alone may struggle to identify the true mechanisms driving a variant’s success especially under the assumption of a fixed growth advantage. By explicitly considering how immunity and transmissibility interact within populations, models that incorporate these dynamics may provide a stronger foundation for understanding why certain variants spread.

### Estimating relative fitness using approximate Gaussian processes

Our earlier approach shows that relative fitness is often dependent on the past exposure of a population (as discussed in Supplementary Text S1 and extended to full immune history models in Supplementary Text S3). This suggests that serology, vaccination history, and immunological data generally can be informative of relative fitness. Additionally, when working with variant classifications, non-neutral evolution within a variant will cause the relative fitness of that variant to change in time. However, even in the absence of external data that can inform relative fitness, there is still hope.

We develop a method for using approximate Gaussian processes to model variant relative fitness. Gaussian processes are probability distributions over functions, where the structure and smoothness of these functions are defined by a kernel that encodes correlations in time. These models are flexible and allow us to encode smoothness constraints, periodicity, and other structures [27]. Gaussian processes allow us a non-parametric estimate of the relative fitness for variants through time (see Materials and Methods).

Traditional Gaussian processes, while flexible, face challenges for large time series and large data sets. Our approach overcomes this using a Hilbert Space Gaussian Process (HSGP) approximation and shared eigenbasis, making the framework scalable for many variants and long time periods [28]. This enables real-time variant fitness estimation and can be applied to any frequency data regardless of the underlying transmission mechanism or immunity assumptions. This model is used in Fig. S2 to estimate the relative fitnesses of different variants through time based on simulated variant sequence counts from frequencies shown in Fig. S1.

Later, we also apply this model to empirical SARS-CoV-2 sequence data from 50 US states and England from 2021 to 2022 to estimate relative fitness for variants circulating in that period.

### Quantifying selective pressure

Although it is useful to quantify the relative fitnesses of individual variants, we are often interested in quantifying the overall effects of selection in the population. With this in mind, we define a population-level metric of overall selective pressure

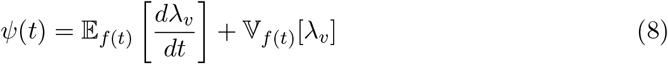

that describes the distribution of relative fitness in the population using the expectation of the fitness change and the variance of fitness. This selective pressure metric serves as an indicator for high fitness variants arising in the population as change. High fitness variants rising from initially low frequency leads to large increases in the variance of the fitness distribution and therefore increases in the selective pressure.

The selective pressure metric enables us to decompose changes in the average growth rate in the population, 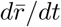, to an evolutionary component *ψ* and a residual baseline growth rate *r*_*W*_ following

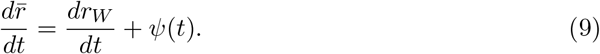

This shows that increased selective pressure through emerging high fitness variants can drive waves of infection. Further, this suggests that differences between growth rates based on selective pressure alone and observed rates are attributable to changes in baseline transmission over time. This mirrors ideas of Fisher’s theorem of natural selection and its later interpretations with the variance of fitness contributing directly to the change in transmission rates (or fitness) [29, 30]. This definition of selective pressure captures how relative fitness contributes to epidemic growth. This is similar to ideas quantifying rates of adaptation via fitness flux [31].

In this case, the overall growth rate 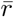 and relative incidence *I*(*t*)*/I*(0) can be written directly

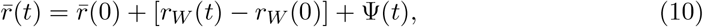

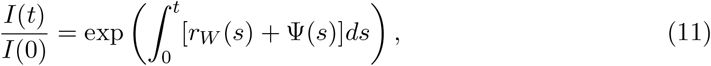

using the cumulative selective pressure 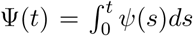. In addition to estimating the relative fitness, metrics derived from these models can inform us of much more.

Our “selective pressure” metric allows us to model the contribution of evolution to changes in the epidemic growth rate of a population and is independent of pivot choice for relative fitness estimation. This metric acts as an early warning system for variant-driven outbreaks, especially in scenarios where case data are sparse or delayed. This metric can be computed using any method that estimates variant frequency and relative fitnesses and serves as a simple tool for understanding the contribution of selection to the overall population dynamics.

The full derivation of this metric and its contribution to the overall growth rate can be found in Supplementary Text S4.

### Predicting epidemic growth rates using selective pressure

Motivated by the relationship between epidemic growth rate and selective pressure demonstrated above, we develop a predictive model of epidemic growth rate using estimates of selective pressure. Using empirical SARS-CoV-2 sequence data from 50 US states between January 2021 and November 2022, we first estimate selective pressure through time using our approximate Gaussian process model on sequence counts (Fig. 4 A–C.) Here, we group variants at the granularity of Nextstrain clades [12] resulting in 28 distinct variants over this time period. As expected we see that relative fitness increases through time and that selective pressure corresponds to speed of clade turnover where the sweep of Omicron BA.1 (clade 21K) yields the strongest signal of selective pressure (Figs. S4–S8). Using case counts from each state, we estimate epidemic growth rates between January 2021 and November 2022. We use these epidemic growth rates to fit a gradient-boosted regressor to predict epidemic growth rates using selective pressure from the most recent 28 days, reserving data between July 2022 and November 2022 for testing (Fig. 4 D–I, Fig. S9). This regressor is chosen via time series cross-validation among model architectures and grid-search parameter tuning (Fig. S10).

**Fig. 4.**
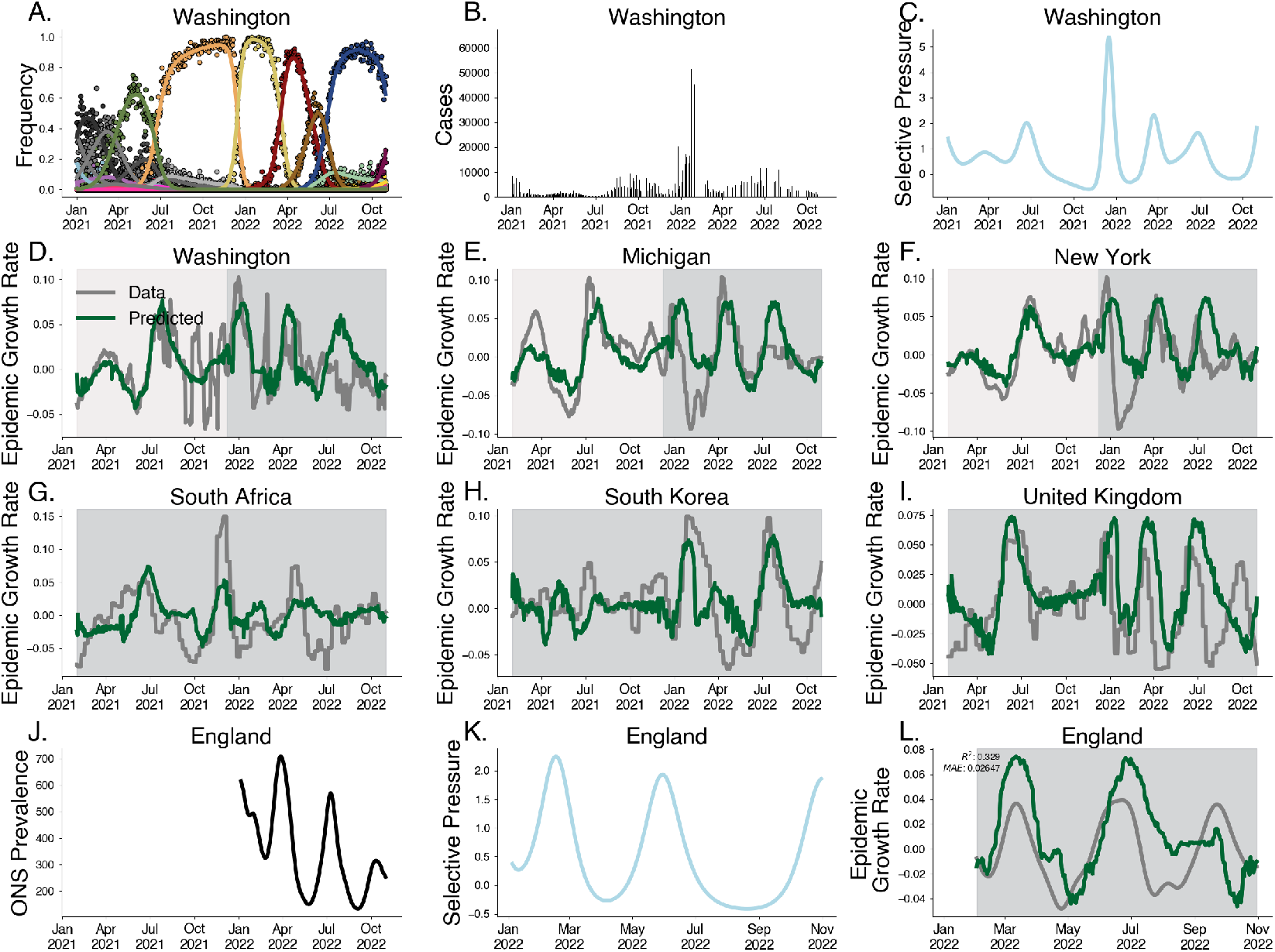
Predicting epidemic growth rate using estimated selective pressure. A. Variant frequency estimated using the Gaussian process relative fitness model between January 2021 and November 2022 for sequence count data from Washington state. B. Case counts from Washington state. C. Selective pressure computed using estimated variant frequencies and relative fitnesses from Washington state. D-F. Predictions for empirical growth rate from selective pressure for selected US states. The light gray period is the training period and the darker gray is the testing period. G-I. Predictions for empirical growth rate from selective pressure for countries South Africa, South Korea and the UK. J. Prevalence estimates for England from ONS Infection Survey. K. Estimated selective pressure in England. L. Empirical growth rates (gray) computed from prevalence estimates and predictions from our model (green) computed from selective pressure.

We observe a strong correspondence between observed epidemic growth rate and model predictions with Pearson *R*^2^ in the training period of 0.576 and a weaker Pearson *R*^2^ in the testing period of 0.077. As case reporting declined over this period, we expect weaker correspondence between our predictions and epidemic growth rates computed from case data. To address this, we sought to evaluate the out-of-sample fit on case data from other countries e.g. South Africa, South Korea, and the United Kingdom, achieving an *R*^2^ of 0.196.

To address the potential for this method under steady reporting rates, we validate this method by predicting the epidemic growth rates in England derived from the Office for National Statistics (ONS) Coronavirus Infection Survey between February 2022 and November 2022. The ONS Infection Survey represented a randomly sampled panel survey of households where nasal swabs were collected regardless of symptom status allowing for prevalence estimates despite faltering case reporting [32]. Our model is able to replicate patterns seen in epidemic growth rates in England derived from ONS data (Fig. 4 J–L), achieving a coefficient of variation of *R*^2^ = 0.329 and mean absolute error of 0.026. Performance is significantly better for the first two subsequent waves, falling off in accuracy for the fall 2022 BQ.1 (clade 22E) wave.

Although these predictions can be biased by non-evolutionary effects on the epidemic growth, this approach provides a simple measure of epidemic growth in the absence of high quality case counts using sequence data alone.

### Latent factor model of relative fitness

The representation of relative fitness using discrete immune backgrounds suggests that there may be low-dimensional structure to variant relative fitness when tranmission is shaped by past host exposure. To elucidate these factors, we develop and implement a method for latent factor analysis of relative fitness from sequence data alone. This model assumes that variants intrinsically escape the immune responses with particular groups and that differences in a variant’s relative fitness between geographies is attributable to differences in immunity between populations. This enables us to estimate pseudo-escape rates for variants as well as pseudo-immunity groups within geographies over time.

We generate Pango lineage-level sequence counts for 18 countries and 53 variants between March 2023 and March 2024. These 18 countries were chosen based on availability of sequence data. Small lineages that do not meet a count threshold are collapsed into their parent lineages. This leaves us with a total of 53 variants, so that each variant met a threshold for number of sequences available.

Using these sequence counts, we apply our latent factor model to estimate the relative fitness of each variant over time in each country, pseudo-escape rates for each variant, and pseudo-immunity for each country simultaneously for *D* = 8 pseudo-immune groups (Fig. 5). This model is significantly constrained relative to estimating the time-varying fitness independently in each location, resulting in a model with 2,752 parameters compared to 7,488 parameters in the independent model. The results of this model are visualized in Fig. 5 for several selected variants and countries of interest.

**Fig. 5.**
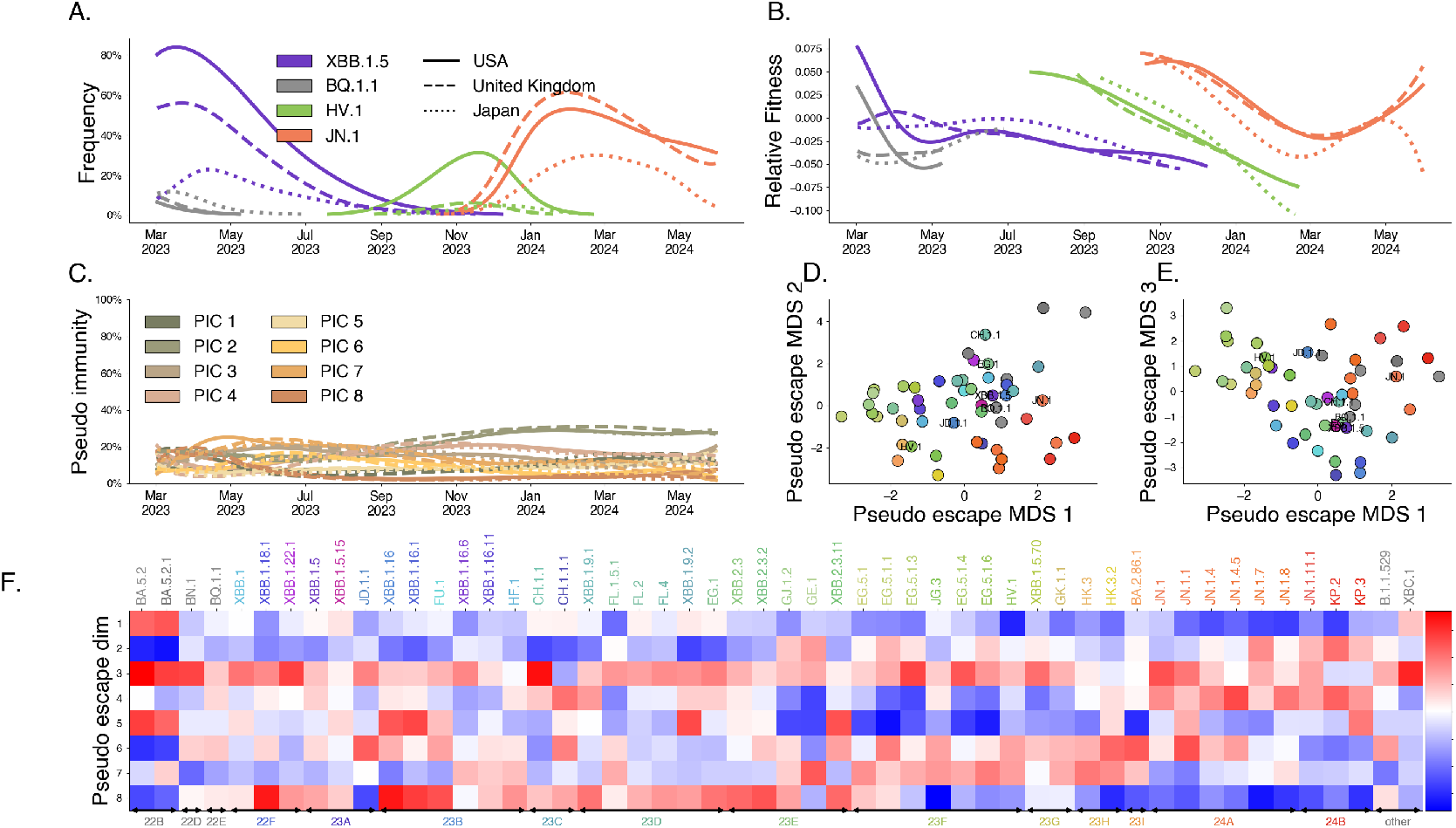
Latent factor models of immunity describe variant dynamics. We fit our latent immunity factor model for *D* = 8 pseudo-immune groups using only SARS-CoV-2 sequence count data. A. Variant frequency. Lines are colored to show 4 variants of interest (of 53 total variants) with the style of the line denoting 3 countries of interest (of 18 total countries). B. Estimated relative fitness for selected variants and countries. These variant-specific relative fitnesses are similar across countries, but not identical. C. Estimated pseudo-immunity cohorts (PIC) over time for multiple countries ordered by decreasing share in the first geography using sequence data alone. D, E. Dimensionality-reduced pseudo-escape rates using multidimensional scaling (MDS). F. Estimated pseudo-escape rates for each variant relative to pivot variant “other”.

We chose *D* = 8 for our primary analysis by noting the point at which the loss function seems to stagnate with increasing *D*, i.e., the “elbow” method (Fig. S11A). Further, we observe that Bayesian Information Criterion (BIC) is minimized between 7 and 9 groups (Fig. S11D). However, the exact choice of latent immune dimensionality is necessarily somewhat arbitrary and we observe significant correlations with empirical titer data for fewer dimensions as well, although *D* = 8 also maximizes this correlation (Fig. S11B) and its significance is maintained for all dimensions *D* > 8 tested. Analogous figures showing pseudo-immunity and pseudo-antigenic relationships across variants can be seen for *D* = 2 in Fig. S12, *D* = 4 in Fig. S13, *D* = 6 in Fig. S14 and *D* = 10 in Fig. S15.

Our results show that closely related Pango lineages are often assigned similar pseudo-escape values. This is visible as a clustering of lineages with similar colors into similar coordinates in Fig. 5D-E suggesting our pseudo-escape values broadly align with evolutionary structure. Further, our model shows that these groups of lineages tend to target particular immune groups such as clade 24A (JN.1, JN.1.1, JN.1.4) has high pseudo-escape in dimensions 3 and 4. If immune escape is the dominant mechanism for relative fitness difference, we expect that differences in immune response between variants from serological data would mirror differences in our pseudo-escape space.

To examine how the learned immune structure relates to subject-level serology, we projected titers from Jian et al. [7] into principal-component space then performed clustering with *k*-means, arriving at *k* = 5 clusters using the elbow method (Fig. S16). Our learned immune clusters yields clear separation (Fig. 6A) in PCA-space, whereas as a subset of subject-level exposure histories show overlap in titer measurements (Fig. 6B). This overlap can be seen in a co-occurrence matrix linking learned clusters to reported exposure histories (Fig. 6C). These groups split and aggregate multiple exposure histories. For example, individuals with a BA.5/BF.7 breakthrough infections split into both clusters 1 and 2, and cluster 3 contains individuals with XBB infection, JN.1 infection, and wildtype vaccine. This indicates that coarse exposure histories may be an imperfect proxy for serological phenotype, and that titer-based clusters may better capture immunological heterogeneity within exposure categories.

**Fig. 6.**
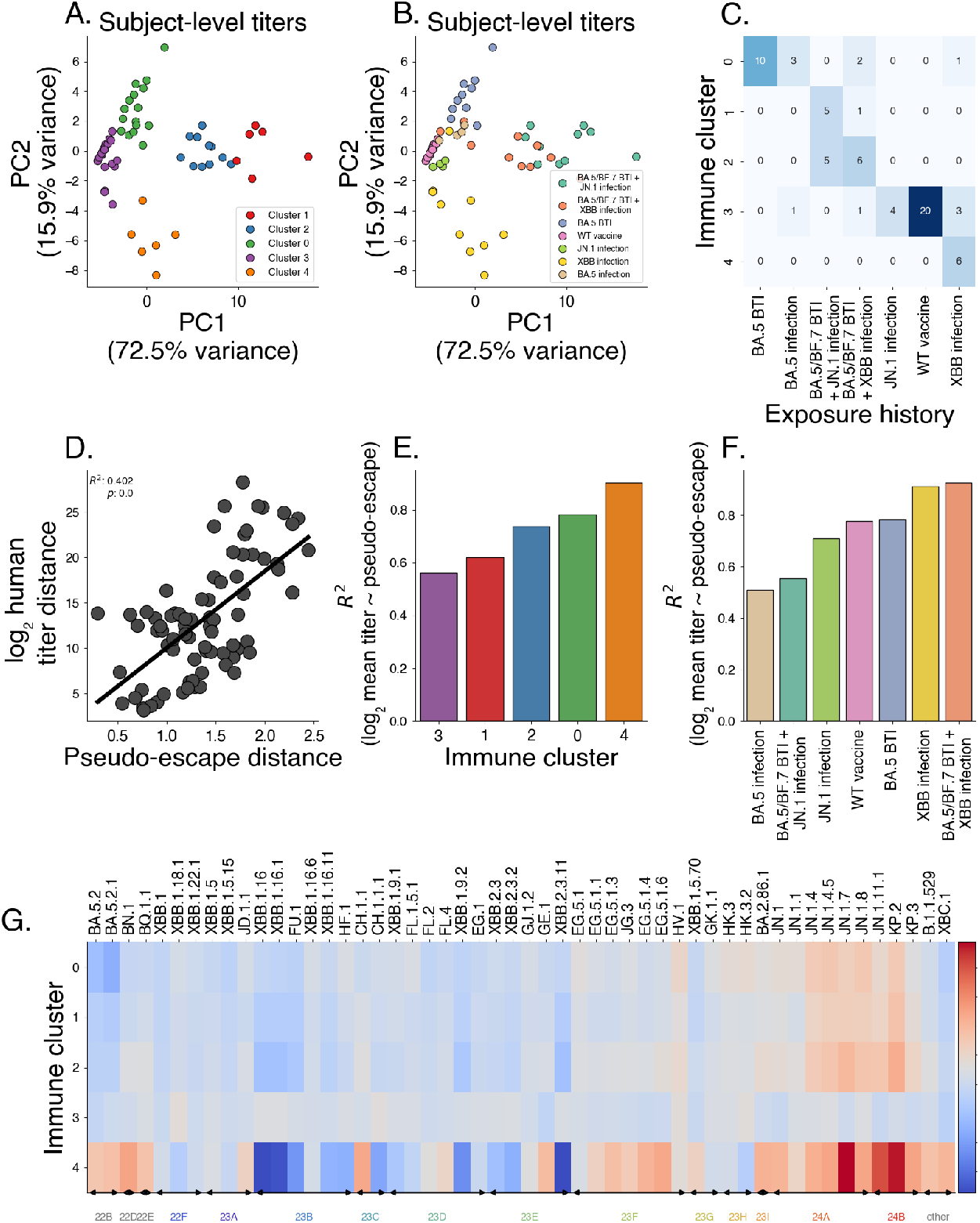
Latent factor models of immunity predict titers across varying exposure histories. Using pseudo-escape values from our latent factor model (Fig. 5) and human titer data, we show that pseudo-escape values predict antigenic distance and titers. (A-B) Principal component analysis of subject-level titers, colored by learned immune clusters (A) and subject infection history (B). (C) Co-occurrence matrix between learned immune clusters and infection histories. (D) Comparing pairwise distance between variants in the pseudo-immune space to observed distances in human titer data. (E-F) Correlation between log mean titer against a variant and that variant’s pseudo-escape by group. (G) Estimated escape burden by immune cluster, showing variant escape potential against individual immune clusters.

We next relate measured serological titers from Jian et al. [7] to our estimates of pseudo-escape that derive solely from variant frequency dynamics across countries. To assess whether pseudo-escape captures titer measurements, we compute serological titer distances as average log2 differences in titer values between pairs of variants. We compare these titer distances to distances in our pseudo-escape space (Fig. 6D), finding the distances between distinct variant pairs in the pseudo-escape space are correlated with these titer differences between variants (*R*^2^ = 0.402). We bootstrap this analysis among 1,000 replicates to assess significance of this relationship (Fig. S17, *p* < 0.001). Next, we subset by exposure history and find a similar relationship in most cohorts with the stronger correlations between serological titer distance and pseudo-immune escape in general being earlier exposures (WT, BA.5) and a weaker correlations observed in cohorts with later exposures (JN.1 and XBB) (Fig. S18).

We quantified which immune clusters are preferentially targeted by circulating lineages using the model-predicted pseudo-escape *η*_*v*,·_ for each variant *v* to predict titers against that variant for individuals in each immune cluster and exposure history group (Fig. 6E-This relationship is delineated in Methods section ‘Regression of pseudo-escape onto neutralization titers’.

To assess variant-level escape against each cluster, we predict an escape burden as the negative of the predicted titer as a proxy for the escape potential against individuals of a certain background. Variants within JN.1 clade show elevated escape burden in specific clusters, with immune cluster 4 standing out as the most broadly targeted. This is consistent with the pseudo-escape patterns in Fig. 5F, where JN.1-family lineages exhibit high pseudo-escape along the dominant dimensions. Further, we find that immune cluster 4 shows a strong negative escape burden again viruses in clade 23B, likely owing the fact that individuals immune cluster 4 corresponds to a subset of individuals with past XBB infection (Fig. 6G). This is further supported by the fact that among all infection histories, pseudo-escape best predicts titers in individuals with past XBB infection including those with BA.5/BF.7 breakthrough infection (Fig. S19-S20).

In short, this shows that a sequence-only latent immunity model can recover an antigenic geometry that agrees with human serology and that learned pseudo-escape values can be used to predict cohort-level neutralization patterns. These properties make the latent representation useful for analyzing population susceptibility and explaining geographic variation in variant success, even when contemporaneous titer data are unavailable.

This approach can be applied to other antigenically variable pathogens, such as seasonal influenza, making it broadly applicable beyond SARS-CoV-2. In fact, there is more utility for pathogens with larger geographic differences in immunity since this approach enables to estimate the proportion of these latent immune pools in the population and how they vary geographically and over time alongside variant difference. By approximating antigenic differences using sequence data alone, this method offers for a deeper understanding of immune dynamics and how they shape variant success in the presence of immune escape. This enables an embedding similar to those from antigenic cartography but without the need for serological data and based purely on observed variant frequencies and estimated variant fitness.

## Discussion

Our study demonstrates the utility of multi-strain mechanistic models in interpreting variant frequency dynamics. This enables a more detailed picture of variant success in environments with heterogeneous population immunity. Our mechanistic grounding of variant fitness allows for investigations into trade-offs between intrinsic transmissibility increase and immune escape, prediction of epidemic dynamics from sequence data alone and inference of antigenic relatedness among variants from differences in success across geographies.

In particular, our latent factor model is most easily compared to the approaches of Meijers et al. [21] and Raharinirina et al. [22] that use cross-neutralization and deep mutational scanning data respectively to parameterize variant fitness. However, our approach differs significantly in that our model does not require any data other than sequence counts for each variant over time, enabling real-time analysis of fitness and heterogeneity in population immunity before cross-neutralization and deep mutational scanning data are available.

Despite these advances, there are limitations to our approach. Long-term forecasts remain difficult, particularly as new variants with unknown fitness profiles emerge. This framework suggests that considering both the escape against individual immune backgrounds and the diversity in human immune escape is most useful for improving forecasts of relative fitness. Our models, while powerful in estimating short-term variant dynamics, rely on assumptions about transmission mechanisms that may not always hold across different pathogens or contexts. In fact, as we’ve shown, it’s entirely possible for shifts in population immunity to change the dominant transmission mechanism.

Furthermore, the models considered here are deterministic in nature and do not explicitly model the emergence of variant viruses only the dynamics after their successful introduction. In reality, there are biological constraints on the types of variants that are produced in nature and even if there is a ‘true’ fitness boost, the chance for stochastic extinction of beneficial variants remains. These constraints present trouble for long-term forecasting as it will require a model of mutation or emergence, tying the potential for a variant to emerge with its potential to transmit in the current environment. Future work should focus on improving the integration of real-time genomic data with serological and epidemiological data, providing a more comprehensive understanding of variant dynamics over time.

In conclusion, our framework represents a significant advance in our understanding of viral evolution and transmission dynamics. By linking variant fitness to specific transmission mechanisms, we provide a more nuanced and accurate prediction of how variants will spread and impact population-level epidemic growth. The selective pressure metric and latent immunity model offer new tools for public health agencies to monitor viral evolution in real time, enabling proactive intervention and insight into the variant difference and wave potential. While our work has been applied to SARS-CoV-2, the methods developed here are broadly applicable to other evolving pathogens, offering a versatile approach for improving epidemic forecasting, variant monitoring, and overall pandemic preparedness.

## Materials and Methods

### Correlations are insufficient for mechanism identification

To assess how vaccination uptake affects the growth advantage of a variant with increased transmissibility, we simulate the spread of a more transmissible variant across populations with different initial past exposure and vaccination levels. This enables us to isolate the effects of transmissibility within different immunity landscapes, examining how relative fitness and growth advantage shift based on population vaccination coverage alone in the absence of immune escape. We begin with the 2-variant SIR model described in Supplementary Text S1. We simulate this model for 100 days with generation time *τ* = 1*/γ* = 3.0 days, *R*_0,*W*_ = 1.4, *I*_*W*_ (0) = 100 individuals, *I*_*v*_(0) = 1 individual, a 50% transmissibility increase *ρ* = 0.5, and no immune escape *η* = 0.0. We divide the period into early and late epidemic with the breakpoint being *t* = 50. In Fig. 3D-E, we estimate the log growth advantage for the variant in the early and full periods using a logit-linear model

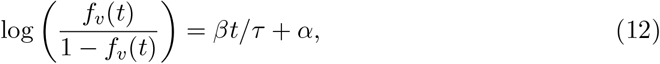

where we take the model slope *β* to be our log growth advantage.

We repeat these simulations for a range of vaccination levels starting from 0% and ending at 65%.

### Generating sequence counts

We prepared sequence count data sets using the Nextstrain-curated SARS-CoV-2 sequence metadata [33] which is created using the GISAID EpiCoV database [34]. These sequences were tallied according to either their annotated Nextstrain clade or Pango lineage [12] depending on the data set to produce sequence count for each variant, for each day over the period of interest, and in each country analyzed.

### Likelihood of sequence counts given frequencies

The models discussed in this paper use observed counts of variant sequences to inform the underlying variant frequency in the population. This is accomplished using a multinomial likelihood, so that given count of sequences *S*_*v*_(*t*) of variant *v* at time *t* and total sequences *N* (*t*) collected at time *t*, we have that

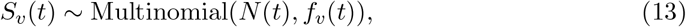

where *f*_*v*_(*t*) is the frequency of variant *v* at time *t*. This is a simple model of sequence counts to frequencies and does not account for over-dispersion of sequence counts relative to a multinomial. However, all models can be extended to estimate and account for over-dispersion by replacing the above likelihood with a Dirichlet-Multinomial likelihood.

### Approximate Gaussian processes for relative fitness estimation

To generate smooth non-parametric estimates of variant growth rates, we develop a Gaussian process based model for relative fitnesses. That is, we model the relative fitness for each variant over time *λ*_*v*_(*t*) as a multivariate normal distribution:

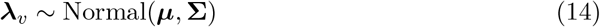

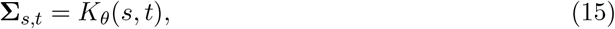

where *K*_*θ*_ is a potentially parameterized kernel function. This induces a structure on the covariance of the relative fitness values over time points *s* and *t*, causing relative fitness to vary smoothly in time.

For computational efficiency, we implement a Hilbert Space Gaussian Process (HSGP) approximation instead of fitting *V* independent Gaussian processes. This approximation allows us to share basis functions between variants [28]. Under this approximation, the relative fitnesses are computed as

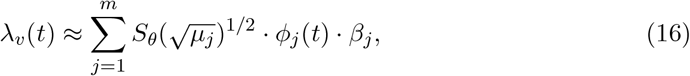

where *S*_*θ*_ is the spectral density of the kernel *K*_*θ*_, *µ*_*j*_ and *ϕ*_*j*_ are the *m* eigenvalues and eigenfunctions of the Laplacian, and *β*_*j*_ ∼ Normal(0, 1) [28]. Since the eigenvalues and eigenfunctions are shared across variants, this allows us to re-use values across variants, simplifying the computation to a matrix multiplication as

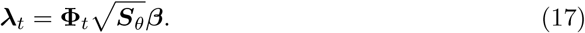

For the analyses in this paper, we use this approximate Gaussian process with a Matérn 5/2 kernel and shared hyperparameters across variants. We demonstrate this model for simulated data from Fig. S1 and show resulting relative fitnesses through time in Fig. S2.

### Predicting epidemic growth rate from selective pressure

The derivation of the selective pressure metric shows that the selective pressure can be a useful tool in predicting the epidemic growth rate. To develop a predictive model of epidemic growth rate using selective pressure, we begin by generating estimates of selective pressure and epidemic growth rate from a period with high sequencing and case surveillance.

We take sequence count and case count data from all states in the United States between January 2021 and November 2022. State-level daily case counts were obtained from USAFacts downloaded on August 7, 2024 at https://usafacts.org/visualizations/coronavirus-covid-19-spread-map/.

Using the sequence counts, we compute selective pressure estimates from relative fitness and frequencies estimated with our approximate Gaussian process relative fitness model. From the case data, we derive the empirical growth rate using a 14-day moving average on case counts *Ĉ*_*t*_ and computing the empirical growth rate as 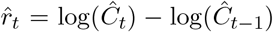. We then use the past 28 days of selective pressure to predict the empirical growth rate.

We use a gradient boosting regressor model which is fit using a mean absolute error loss function. This model was selected as it achieved the minimal error via time series crossvalidation averaged across 10 splits among candidate models (Fig. S10). The candidate models include linear regression, ridge regression, Lasso regression, random forests, and gradient-boosted trees as implemented in scikit-learn [35]. We additionally tune the hyperparameters of this model using grid search cross-validation.

We validate our model by comparing our predicted epidemic growth rates to held-out case data for US states, South Africa, South Korea, the United Kingdom, and additionally to estimates of the epidemic growth rates in England derived from data from the Office for National Statistics (ONS) Coronavirus Infection Survey [32]. Estimates of prevalence from the ONS Infection Survey were obtained for January 2022 to September 2022 from www.ons.gov.uk/peoplepopulationandcommunity/healthandsocialcare/conditionsanddiseases/datasets/coron Epidemic growth rates are computed on this data in the same way as the state-level analysis.

### Latent immune factor model

We show that relative fitness dynamics can be explained by low-dimensional immunity when transmission dynamics are described with compartmental models (Supplementary Text S1). This motivates a model to learn this low-dimensional structure that is inspired by latent-factor models. We start by assuming that the relative fitness of variant *v* at time *t* and in geographic location *g* can be described by *D* latent factors so that

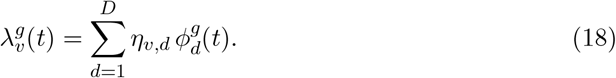

As the structure here resembles Equation 43, we call *η*_*v,d*_ “pseudo-escape” of variant *v* from group *d* and 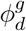 “pseudo-immunity” group *d* in geographic location *g*. To make this more consistent with our intuition here, we model 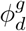 to be in [0, 1] and model it as smoothly varying in time. We model logit 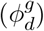 using 4th order splines with 6 knots placed uniformly over the time period modeled. Though we choose to model these latent factors with splines, other models would work here. For example, one alternative would be the approximate Gaussian processes described above. Additionally, in order to ensure identifiability of the parameter estimates, we fix some base variant *v*^∗^ which fitness is defined relative to, so that 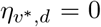 for all 1 ≤ *d* ≤ *D*. For the same reason, we fix the order of components, so that the components are numbered in decreasing order by their share in the arbitrarily defined base geography.

We apply this model to SARS-CoV-2 sequence counts in the period between March 2023 to March 2024 for 14 countries. To access the necessary number of immune dimensions, we vary the number of immune dimensions between *D* = 2 to *D* = 12. Looking at the loss for the latent factor model for increasing *D*, we choose *D* = 8 for our primary analysis by noting the point at which the loss function seems to stagnate with increasing *D* i.e. the “elbow” method (Fig. S11).

We compare the distances between variant pairs in our estimated pseudo-escape space to distances in log2 titer. Using human titer data from Jian et al [7], we compute neutralization titer distances as the average of differences in log2 neutralization titers between pairs of variants for a cohort of individuals. This analysis is repeated among 1,000 bootstrapped samples to create a distribution of *R*^2^ values (Fig. S17). Additionally, we subset this by exposure history and repeat this analysis to find which exposure groups best explain distances in pseudo-escape space (Fig. S18).

### Regression of pseudo-escape onto neutralization titers

To assess the relationship between our estimated pseudo-escape values and empirical neutralization titers, we use human titer data from Jian et al. [7]. For each exposure group, we have a set of neutralization titers measured against multiple variants. Our goal is to determine whether variation in titers across variants can be explained by the pseudo-escape values inferred from our latent immune factor model.

Let *t*_*i,v*_ denote the neutralization titer of individual *i* against variant *v*. For each exposure group *G*, we first aggregate titers by computing the group-level mean and applying a log_2_ transformation with a pseudocount of one, i.e.

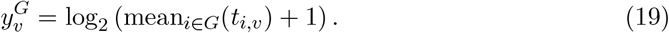

Let *η*_*v*_ ∈ ℝ^*D*^ denote the pseudo-escape vector of variant *v* estimated from the latent immune factor model across *D* latent factors. We then fit an ordinary least squares (OLS) model separately for each exposure group,

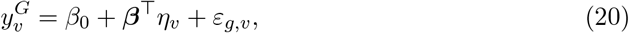

where *β*_0_ is an intercept term and ***β*** is a vector of regression coefficients mapping latent factors to predicted group-level titers. This yields a coefficient of determination 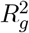 for each group, quantifying how well variation in pseudo-escape explains variation in group-level titers.

To assess statistical significance, we performed a permutation test in which the association between variants and their pseudo-escape vectors was randomly permuted 1,000 times. For each permutation, we re-fit the OLS model and recorded the resulting 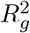, yielding a null distribution under the hypothesis that pseudo-escape and titers are unrelated. The empirical *p*-value for each group was computed as the fraction of permutations with 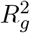 greater than or equal to the observed 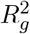. This procedure controls for the correlation structure in the titer data while directly testing whether the inferred pseudo-escape values have predictive power for neutralization measurements.

We summarize the results across groups by reporting both the observed *R*^2^ values and the permutation-derived *p*-values (Fig. S19 and S20). This analysis identifies exposure groups where pseudo-escape provides a statistically significant explanation of the neutralization response profile.

## Data Availability

Source code used to generate figures, model implementations, and sequence count data are available at github.com/blab/relative-fitness-mechanisms.

## Acknowledgements

We thank Ivana Bozic, Betz Halloran, Mark Kot and Erick Matsen, as well as members of the Bedford Lab for their feedback on this work. We gratefully acknowledge all data contributors, ie the Authors and their Originating laboratories responsible for obtaining the specimens, and their Submitting laboratories for generating the genetic sequence and metadata and sharing via the GISAID Initiative, on which this research is based. We have included an acknowledgements table in the associated GitHub repository under data/final acknowledgements gisaid.tsv.xz.

## Author contributions

MF conceived the study. MF, TB gathered sequence and case count data. MF designed and implemented the models. MF performed the analysis. MF, TB interpreted the results. MF, TB wrote the paper.

## Supplementary Information

## Supplementary Text

### S1 Exponentially growing populations to frequency dynamics

We consider a viral population consisting of *V* exponentially-growing variant viruses each with prevalence *I*_*v*_. Defining the time-varying growth rate for the prevalence of variant *v* as *r*_*v*_(*t*), we can model the prevalence using an ordinary differential equation

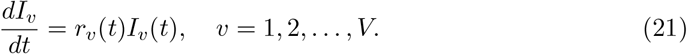

The above differential equation has a known solution in terms of the integral of the time-varying growth rate and initial prevalence,

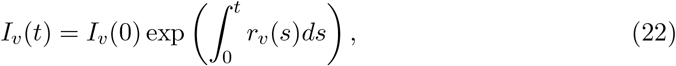

where *I*_*v*_(0) is the initial prevalence of variant *v*.

Now turning to the frequency dynamics of the population, we write the frequency of variant *v* in the population as 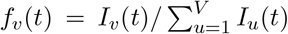. This allows us to derive an ODE for variant frequency in terms of the variant growth rates using the quotient rule for differentiation

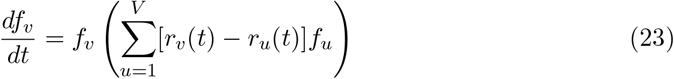

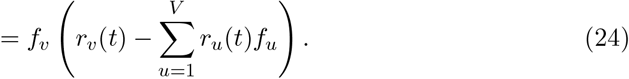

This system of differential equations resembles a logistic growth equation and can be shown to have the following solution in terms of the initial frequencies *f*_*v*_(0) and the variant growth rates

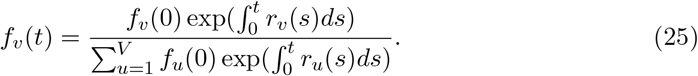

The above representation of the variant frequency will serve as a centerpiece for many of the arguments to follow. We see that by tracking the rate at which variant viruses are spreading, we can construct the corresponding frequency dynamics without knowing the absolute prevalence of any variant.

#### Relative frequency and relative fitness

Using the above equation for the variant frequencies, we can write the relative frequency of variant *v* over *u* as *x*_*v,u*_(*t*) = *f*_*v*_(*t*)*/f*_*u*_(*t*) to see

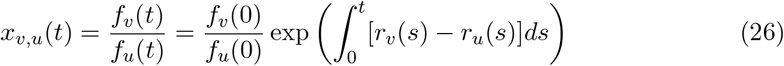

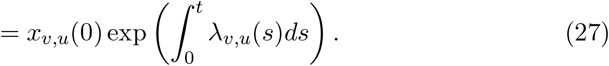

Notice this relative frequency change depends on the initial relative frequencies and the *relative fitness λ*_*v,u*_(*t*) = *r*_*v*_(*t*) − *r*_*u*_(*t*) of *v* over *u*. This relative fitness has the same units as the exponential growth rate (e.g. per day). Using the definition of relative fitness, we can notice that

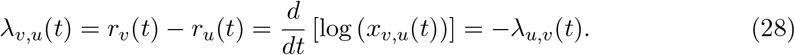

We can see that there is a symmetry in the relative fitnesses and that the associated frequency dynamics depend on the differences between relative fitnesses. This suggests that absolute fitness (in terms of the growth of infections) may not be inferable from frequencies alone. This definition of relative fitness becomes essential in describing various existing modeling approaches for frequency dynamic data and motivates possible extensions since we can represent these models as having the form:

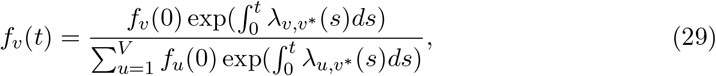

where the relative fitness of *v* is expressed as relative to an arbitrary pivot variant *v*^∗^.

#### Cumulative relative-fitness and frequency change

Above we saw that within our framework frequency change over time intervals depends only on the cumulative relative fitness over time intervals 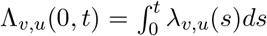. We can then characterize approaches for modeling frequency change in terms of how they represent, estimate, and forecast these relative fitnesses. This framework includes various existing methods for analyzing frequency data such as the seasonal influenza forecasting models of Lässig and Luksza [36] and Huddleston et al [37], multinomial logistic regression for frequency estimation [13] and the SARS-CoV-2 mutational fitness model of Obermeyer et al [38].

Though this framework can be used to describe existing statistical methods for frequency modeling, it is also applicable to traditional compartmental models of epidemics. In fact, applying these ideas to compartmental models enables to see how mechanistic assumptions on the transmission process determine relative fitness of variant viruses.

#### Two-strain SIR

For simplicity, we will begin by analyzing a two-strain SIR model in which a variant virus *v* can differ from wildtype virus wt by increased intrinsic transmissibility (via *η*_*T*_) and immune escape against wild-type immunity (via *η*_*E*_). This gives a system of 5 ordinary differential equations

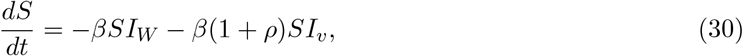

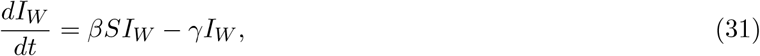

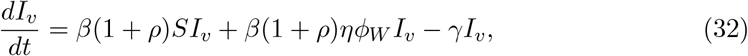

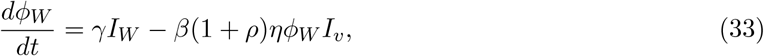

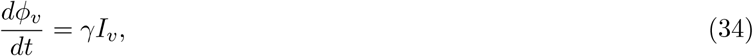

where *I*_*W*_ denotes wild-type prevalence, *I*_*v*_ denotes variant prevalence and *ϕ*_*W*_ denotes immunity derived from wild-type infection. In this model, the variant virus can infect both susceptible individuals *S* and individuals with immunity to wild-type virus *ϕ*_*W*_ . Increased intrinsic transmissibility increases the baseline transmission rate from *β* in wild-type to *β*(1 + *ρ*) in the variant virus and immune escape increases the transmission rate against those with wildtype immunity, so that the at-risk population is *ηϕ*_*W*_ .

Writing that *r*_*W*_ (*t*) = *βS* − *γ* and *r*_*v*_(*t*) = *ρβS* + *β*(1 + *ρ*)*ηϕ*_*W*_ − *γ*, we can then write the relative fitnesses as:

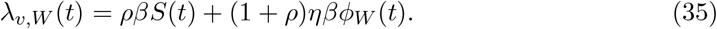

From this representation of relative fitness, we can see that given fixed increases to overall transmission (*ρ* > 0) or immune escape (*η* > 0), the observed fitness boost at the level of variant relative fitness still depends on the proportion of the population at risk for infection.

#### *n*-strain SIR

This model can also be extended to an *n*-strain SIR model where each variant strain *v*_*i*_ with 2 ≤ *i* ≤ *n* is described by its own advantage parameters *θ*_*i*_ = (*ρ*^(*i*)^, *η*^(*i*)^) relative to the wildtype (*θ*_*W*_ = *θ*_1_ = (0, 0))

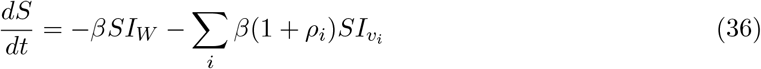

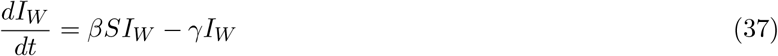

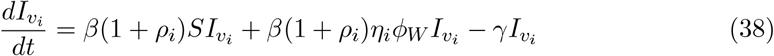

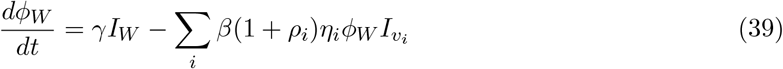

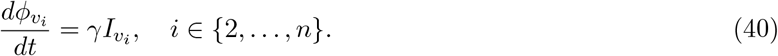

In this formulation, the variant viruses compete only for susceptible population and those with previous wild-type infection. This formulation can be generalized to allow for competition between all variants for any exposure history and will be discussed in the following sections. In Fig. S1, we implement and simulate a 3-strain model with wildtype as above, an escape variant E with *θ*_2_ = (0, *η*), and a transmissibility increase variant T with *θ*_3_ = (*ρ*, 0).

#### Models of immune escape against heterogeneous backgrounds

We’ll now consider a model where all hosts are assumed to fall into one of *B* immune backgrounds *ϕ*_*b*_ for *b* = 1, … , *B*. We assume that infection by each variant *v* then leaves recovered hosts in the corresponding immune background of the most recent infection *b*_*v*_. Variant transmission then occurs via immune escape against a background leading to a matrix of escape rates ***η*** = *η*_*v,b*_ for variants *v* and background *b*.

We can then write the system of ordinary differential equations as

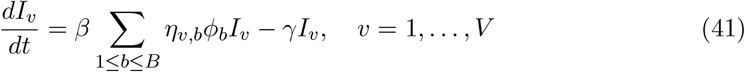

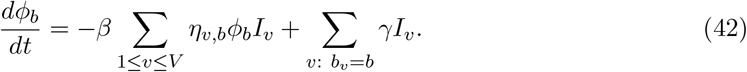

With this model, susceptible and recovered compartments in the standard SIR model can be thought of as immune backgrounds. This allows us to represent the standard SIR model as *S* = *ϕ*_*S*_, *I* = *I*_*W*_ , *R* = *ϕ*_*W*_ and *η*_*W,S*_ = 1, *η*_*W,W*_ = 0 and *b*_*W*_ = *W* . We can also think of the two-strain SIR with *ρ* = 1 as a special case of this model where we set *S* = *ϕ*_*S*_, *η*_*W,S*_ = 1, *η*_*W,W*_ = 0, *η*_*v,S*_ = 1, *η*_*v,W*_ = *η* and keep all other parameters the same.

With this formulation of immune escape, we can then write the relative fitnesses in terms of the escape rates *η*_*v,b*_ and the immune background proportions *ϕ*_*b*_ as

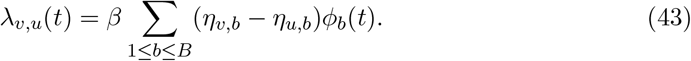

Under this model of immune escape, we can see relative fitness among variants can be decomposed into differences in immune escape among immune backgrounds within a population. Due to the dependence here on the proportion of each immune background in determining fitness, this suggests that the overall distribution of susceptibility to strains is potentially an important consideration when translating individual-level measures of immune escape to population-level estimates of variant fitness. Understanding the size and complexity of this immune space may therefore be useful for parameterization and forecasting of variant frequencies. However, the extent to which modeling this complexity affects estimates of relative fitness also depends on how quickly the distribution of immune backgrounds change i.e. 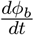.

Though the derivation above uses a simplified model of using most recent infection to sort individuals into an immune group, we show that a more complicated model that accounts for the entire exposure history of the host also gives a similar decomposition to relative fitness in Supplementary Text S3.

### S2 Revisiting existing models for frequency growth

Using the theory developed for exponentially-growing variant populations, we now re-visit existing methods for modeling viral frequency dynamics.

#### Multinomial Logistic Regression

We begin with multinomial logistic regression (MLR) with fixed relative fitness. This model can be written as

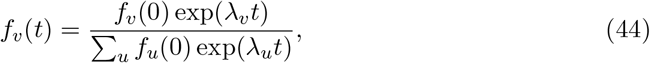

where *f*_*v*_(*t*) is the frequency of variant *v* at time *t* and *λ*_*v*_ is the relative fitness of variant *v*. This provides estimates of the relative fitness compared to some reference strain *u*^∗^ for which 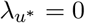. In this model, initial frequencies *f*_*v*_(0) and relative fitness *λ*_*v*_ are estimated from frequency dynamics. Converting this estimate to an estimate of transmission advantage (relative effective reproduction number) requires assuming a Dirac delta distribution of the generation time [39].

Comparing this to equation 35, we can see this model of fixed relative fitness results from assuming that the at-risk populations are constant over-time. This assumption is useful since it requires no outside knowledge of the at-risk population and relative infection rates, though this may be less useful for longer forecasts or when there is large turnover in at-risk populations due to infection.

#### Fitness models of seasonal influenza

Motivated by the observed antigenic evolution of seasonal influenza, Lässig and L-uksza [36] and Huddleston et al [37] approximate the cumulative relative fitness between influenza seasons on the level of individual strains as

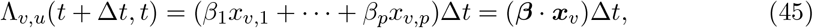

where the relative fitness is determined by strain-specific predictors ***x***_*v*_ and the regression parameter ***β***_*v*_ are estimated.

This formulation fits neatly into the framework we’ve developed as the cumulative fitness here can be written as the integral of a relative fitness *λ*_*v,u*_ = ***β*** · ***x***_*v*_ over the time period of interest:

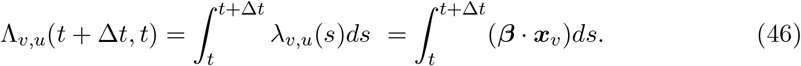

Therefore, these models can be thought as regression-based predictors of relative fitness where frequency and external covariates contribute to estimated relative fitness.

### S3 Relative fitness for full immune history models

We show that the simple background model is consistent with an expanded immune history model. Beginning with the model from Lazebnik and Bunimovich-Mendrazitsky 2022 [40], we consider the differential equation for the individuals with strain infection history *J* and current infecting strain *i R*_*J*_ *I*_*i*_

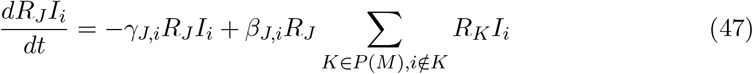

where *P* (*M*) is the collection of all finite infection histories.

Here, infection can occur from any individual infected with strain *i* assuming their past immune history does not include *i* and the infected are any recovered individual with immune history *J R*_*J*_ . To compute the strain growth rate, we can sum over all possible immune histories for individuals infected with strain *i*, so that

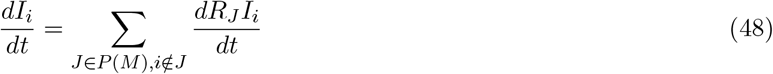

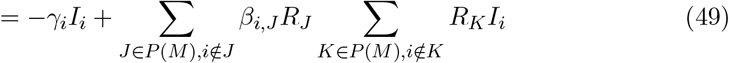

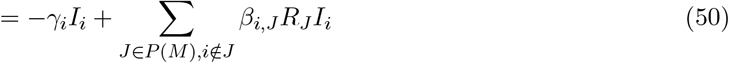

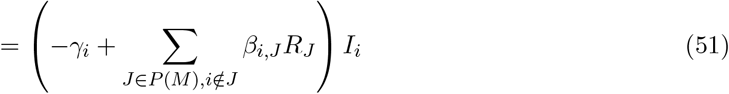

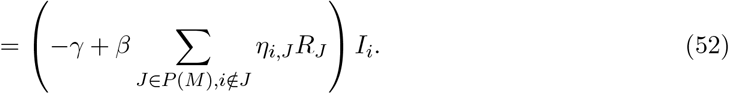

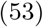

Assuming that the transmission rate can be decomposed as a base transmission rate *β* and a strain *i* and immune history *J* specific escape rate *η*_*i,J*_ and that the recovery rate is constant, we notice this is identical to our previous immune background model. Therefore, our relative fitnesses simplify to

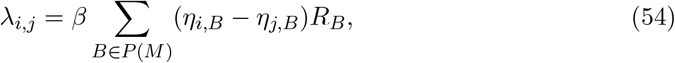

where for simplicity we define *η*_*v,B*_ = 0 if *v* ∈ *B*.

### S4 Selective pressure and contribution to epidemic growth rates

In this section, we derive our selective pressure metric *ψ*(*t*) and show how it contributes to the overall epidemic growth rate in the population.

Beginning again from our assumption of inhomogeneous exponential growth, we can write a differential equation for the total prevalence *I*(*t*) = ∑_*v*_ *I*_*v*_(*t*),

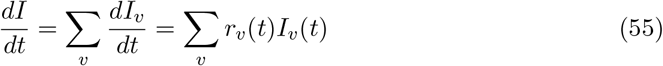

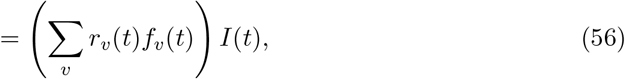

where we’ve used that *I*_*v*_(*t*) = *f*_*v*_(*t*)*I*(*t*). This allows us to see that 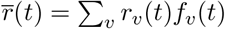 is the average growth rate of the prevalence. Re-writing the average in terms of some base exponential growth rate and the relative fitnesses so that *r*_*v*_(*t*) = *λ*_*v*_(*t*) + *r*_*W*_ (*t*), we get that 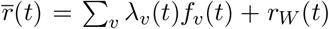. We can simplify this by writing 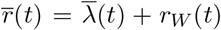 where 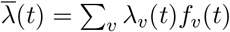 is the mean fitness of the population. We can now look at the rate of change in the average growth rate by taking its derivative

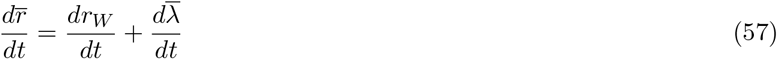

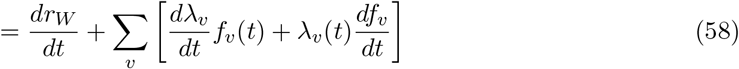

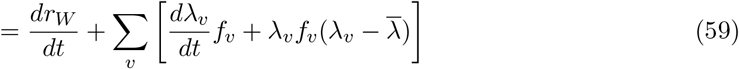

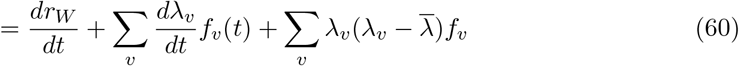

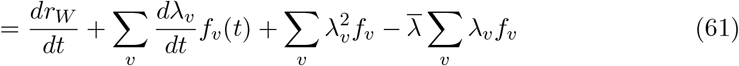

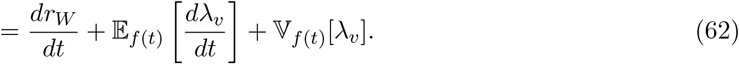

Here, we’ve written the last line in terms of expectations relative to sampling according to the frequency distribution. This shows us that the change in the average growth rate of the epidemic can be written in terms of the growth rate of the pivot category, the mean rate of change in the relative fitness, and the variance of the relative fitnesses. We will call terms which can be computed in terms of quantities derived from frequencies alone the selective pressure

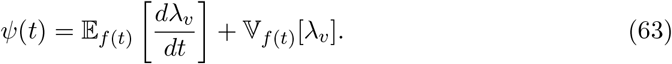

We can use this idea to directly write the prevalence in terms of the selective pressure and the base growth rate. First, we define a cumulative selective pressure

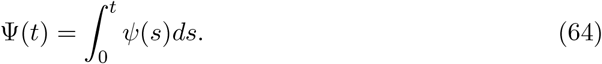

We can then use this to reconstruct the relative incidence

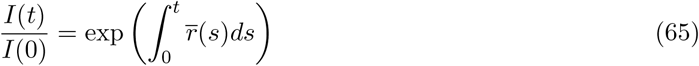

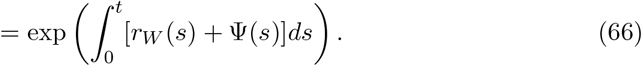

## Supplementary Figures

**Fig. S1.**
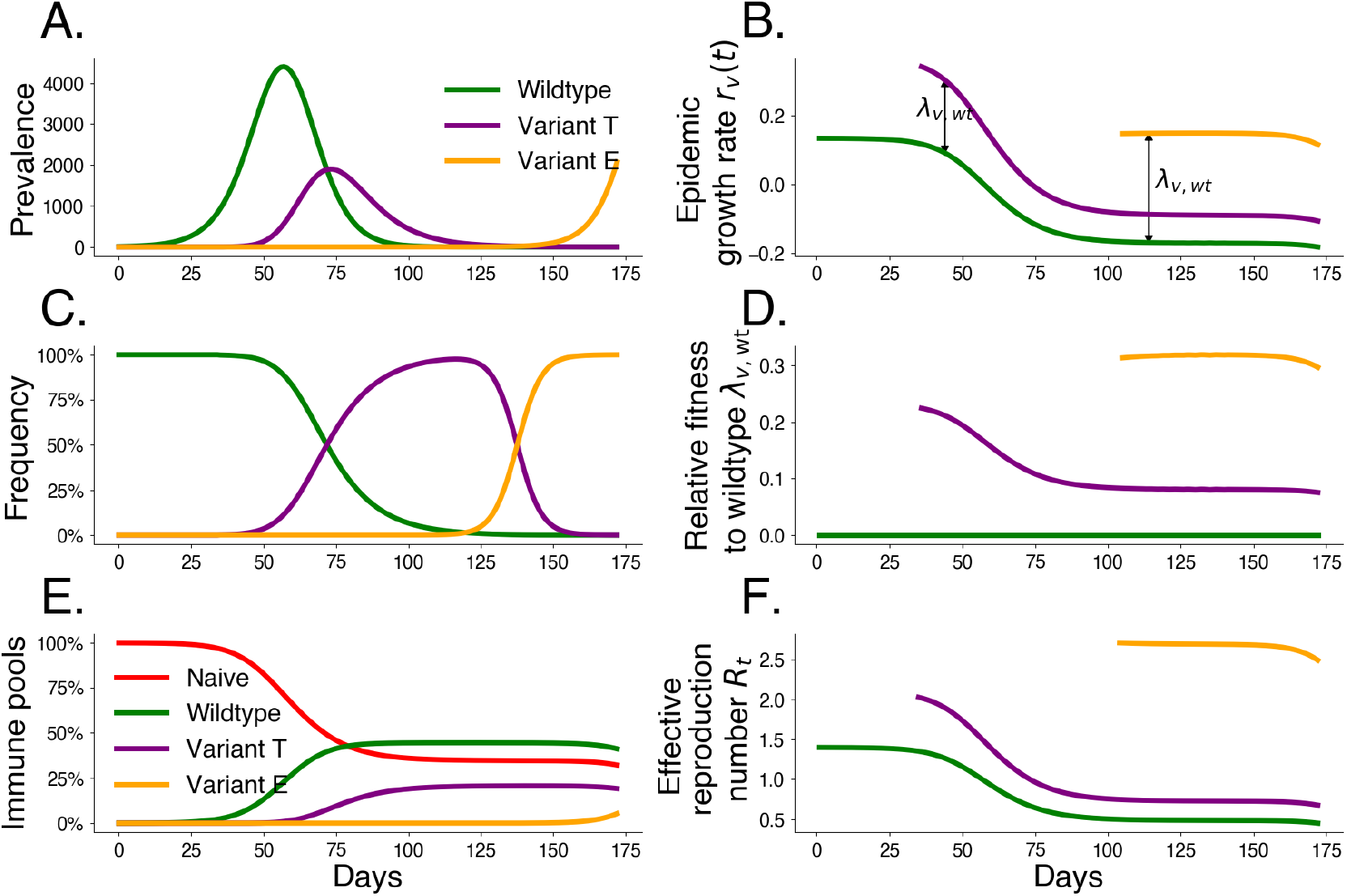
Simulated variant dynamics in a mechanistic model. Mechanistic transmission models constrain variant frequency dynamics by specifying a functional form for relative fitnesses. Simulations of a three-variant model including wildtype *W* , an intrinsic transmission variant *T* , and an immune escape variant *E* show the relationship between population-level transmission and selection. We begin the simulation with initial wildtype prevalence *I*_*W*_ (0) = 1, effective reproduction number *R*_0,*W*_ = 1.4, and duration of infection 1*/γ* = 3.0 days. We introduce transmissibility variant *T* at *t* = 20 with frequency *f*_*T*_ (20) = 10^−5^ and a 50% increase in transmissibility *ρ* = *ρ*_*T*_ = 0.5. We introduce escape variant *E* at *t* = 70 with frequency *f*_*E*_(70) = 10^−6^ that infects 5% of hosts possessing wildtype immunity *η* = *η*_*E*_ = 0.05. A. Prevalence *I* by variant. B. Exponential growth rate *r* by variant. C. Variant frequency *f* . D. Fitness relative to wildtype *λ*. E. Underlying immune pools. F. Effective reproduction number *R*_*t*_ by variant.

**Fig. S2.**
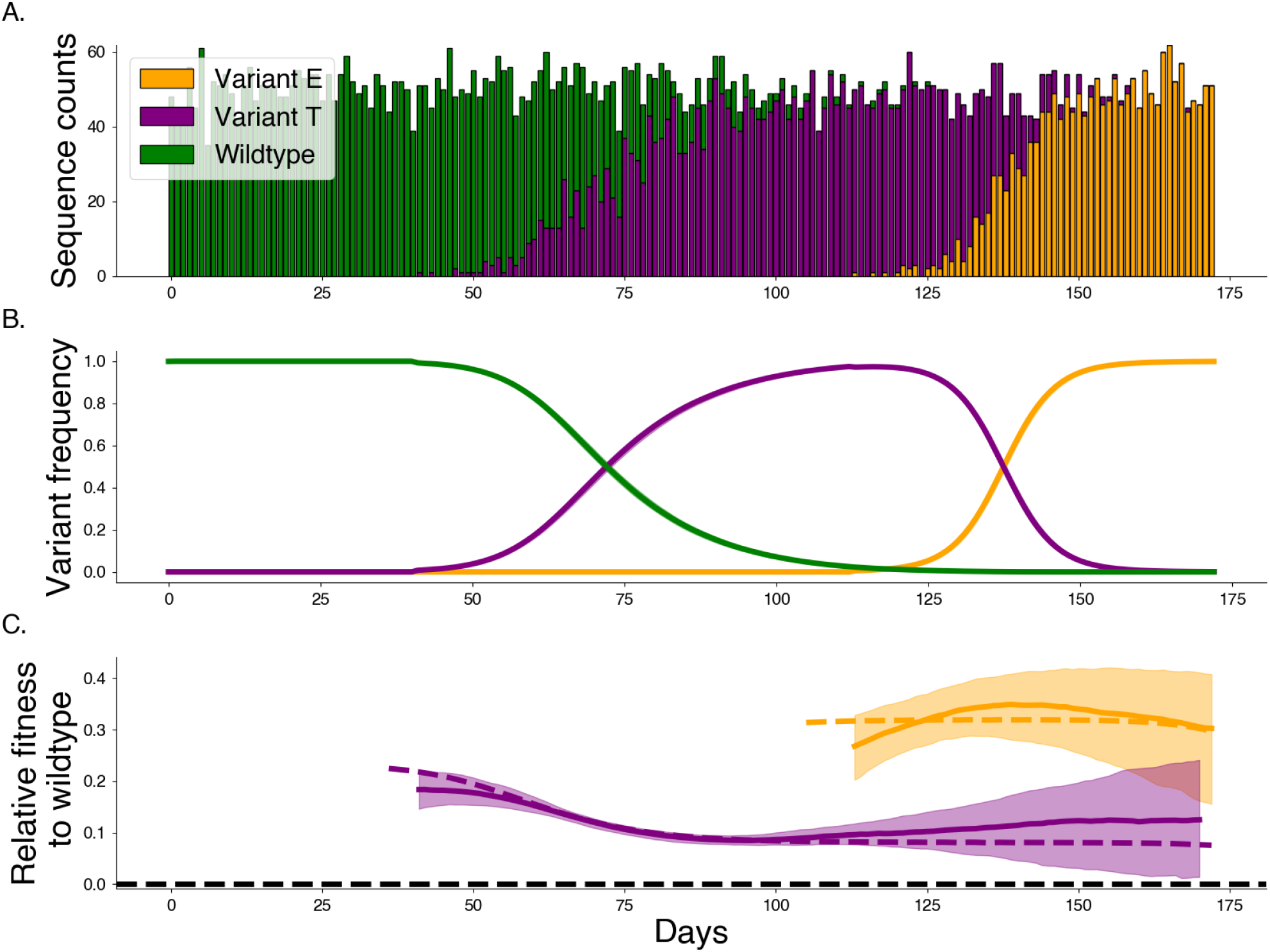
Estimating relative fitness with Gaussian processes. Gaussian processes allow us a non-parametric estimate of the relative fitness for variants through time. This figure uses Gaussian processes to model the 3 variant example shown in Fig. S1. A. Synthetic sequence counts generated using a multinomial distribution with frequencies from Fig. S1C. B. Frequencies and posterior frequencies according to Gaussian process model. Intervals show the 80% credible interval. C. Posterior relative fitnesses. Intervals show the 80% credible interval. Dashed line shows true relative fitnesses from underlying mechanistic model.

**Fig. S3.**
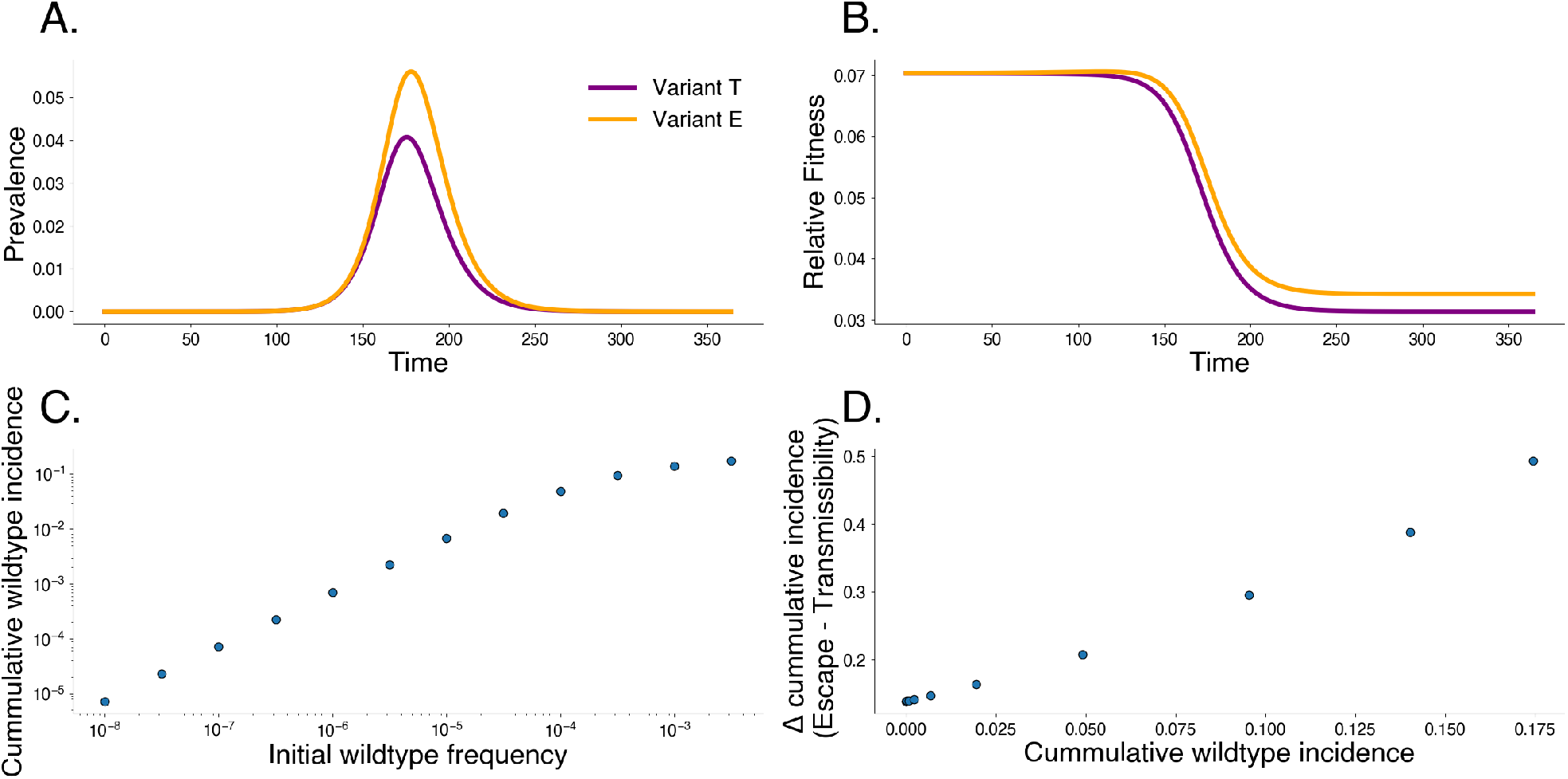
Differences in fitness mechanisms impact frequency and prevalence in the short-term. Comparing simulations from two independent two-variant systems with either an escape variant E (orange) or a transmissibility variant T (purple). We fix the initial relative fitness for the two variants using Equation 4 and simulate dynamics for 365 days. A. The prevalence for the variants. B. The relative fitness from the variants. C. The cumulative wildtype incidence as a function of the initial wildtype frequency. D. The difference between the cumulative incidence between the escape variant and the transmissibility variant as a function of wildtype incidence.

**Fig. S4.**
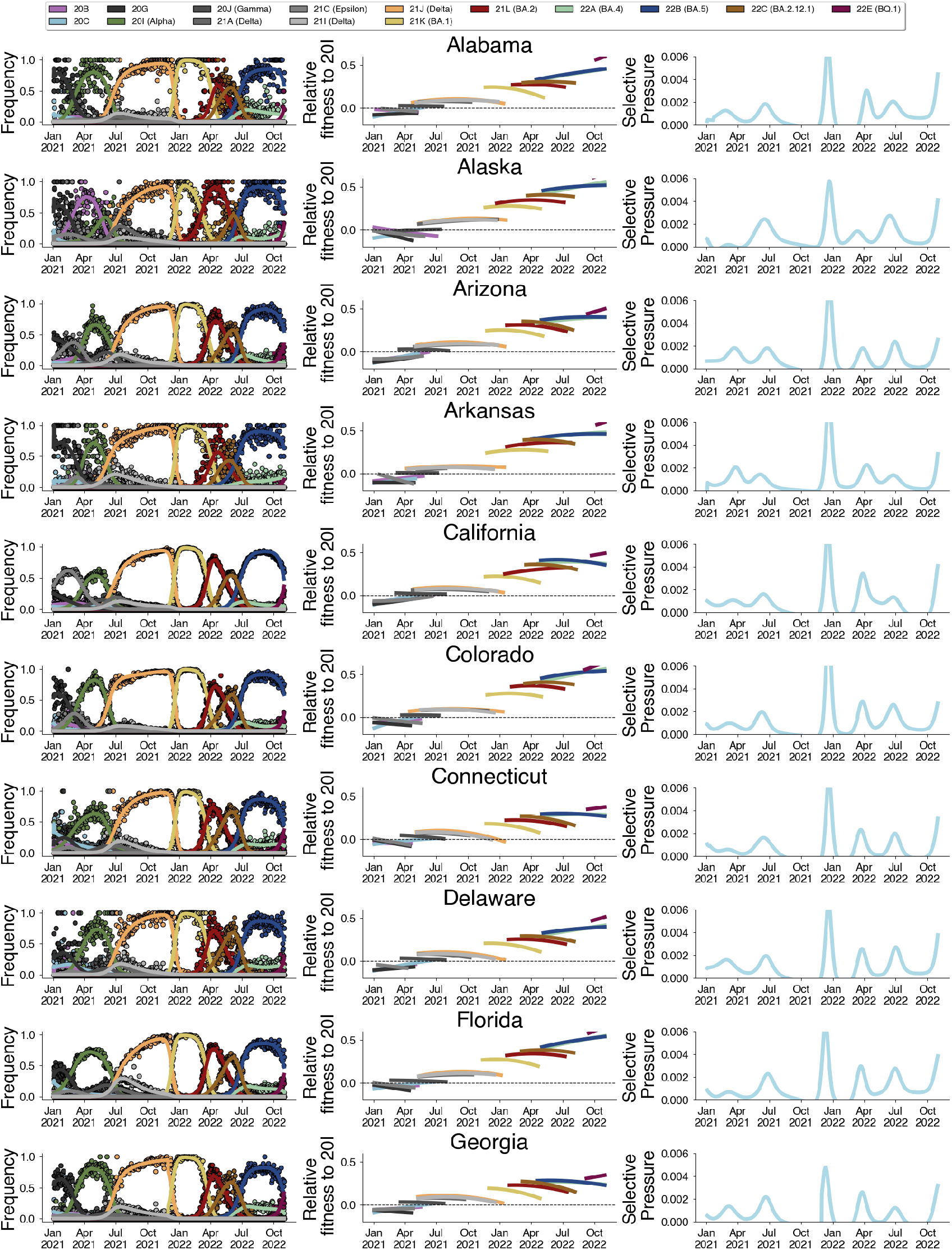
Estimated variant frequencies, relative fitnesses, and selective pressure. Alabama through Georgia.

**Fig. S5.**
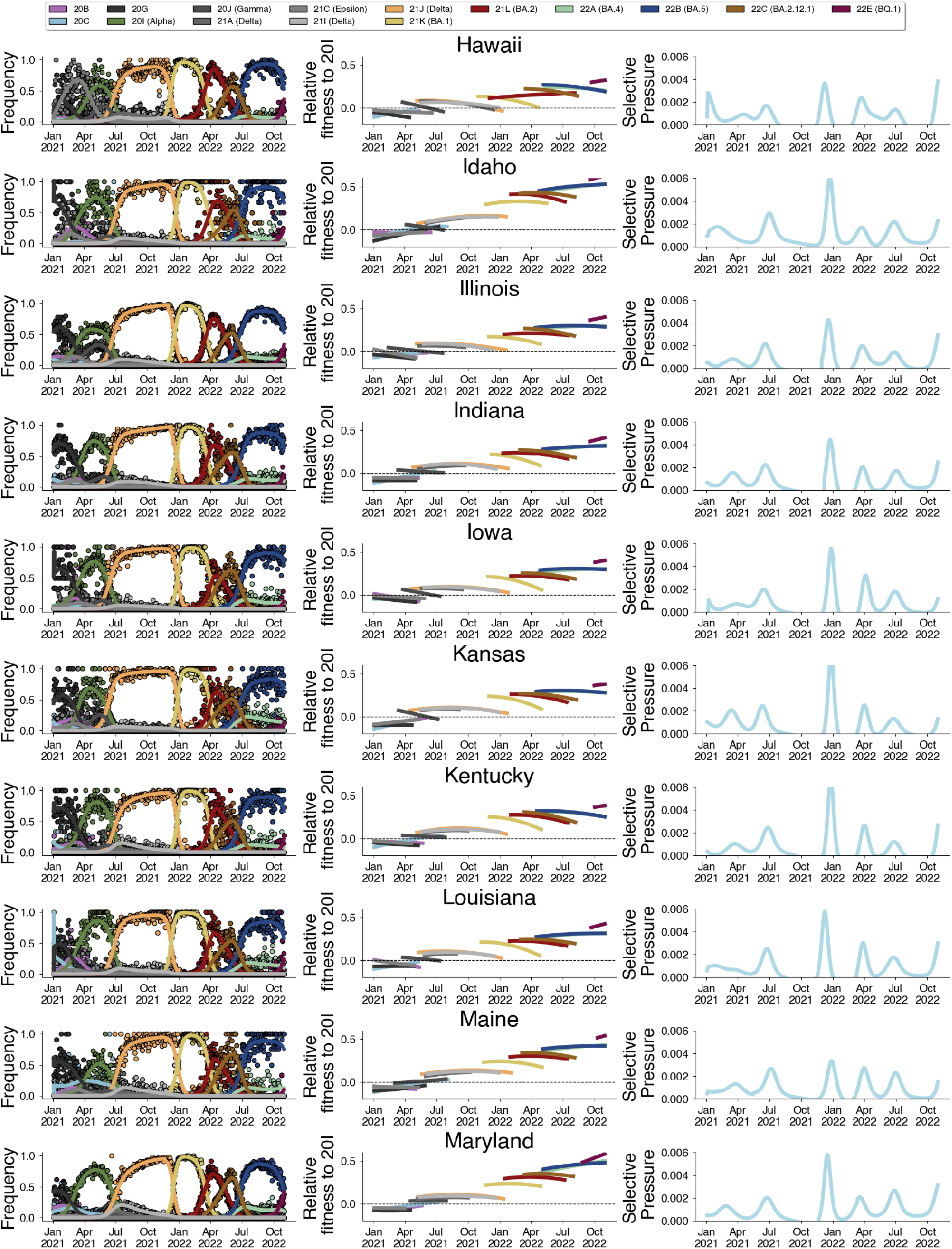
Estimated variant frequencies, relative fitnesses, and selective pressure. Hawaii to Maryland.

**Fig. S6.**
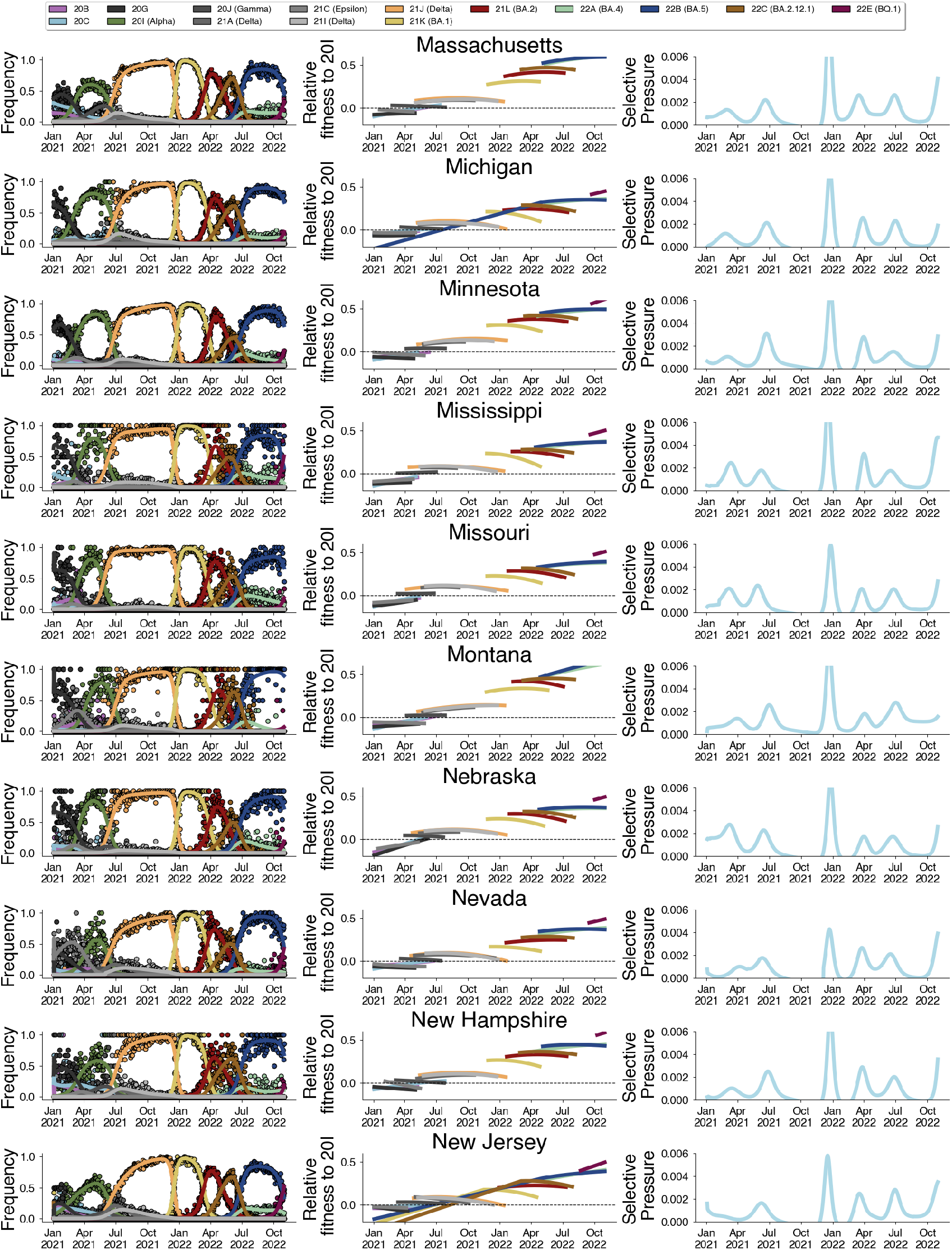
Estimated variant frequencies, relative fitnesses, and selective pressure. Massachusetts to New Jersey.

**Fig. S7.**
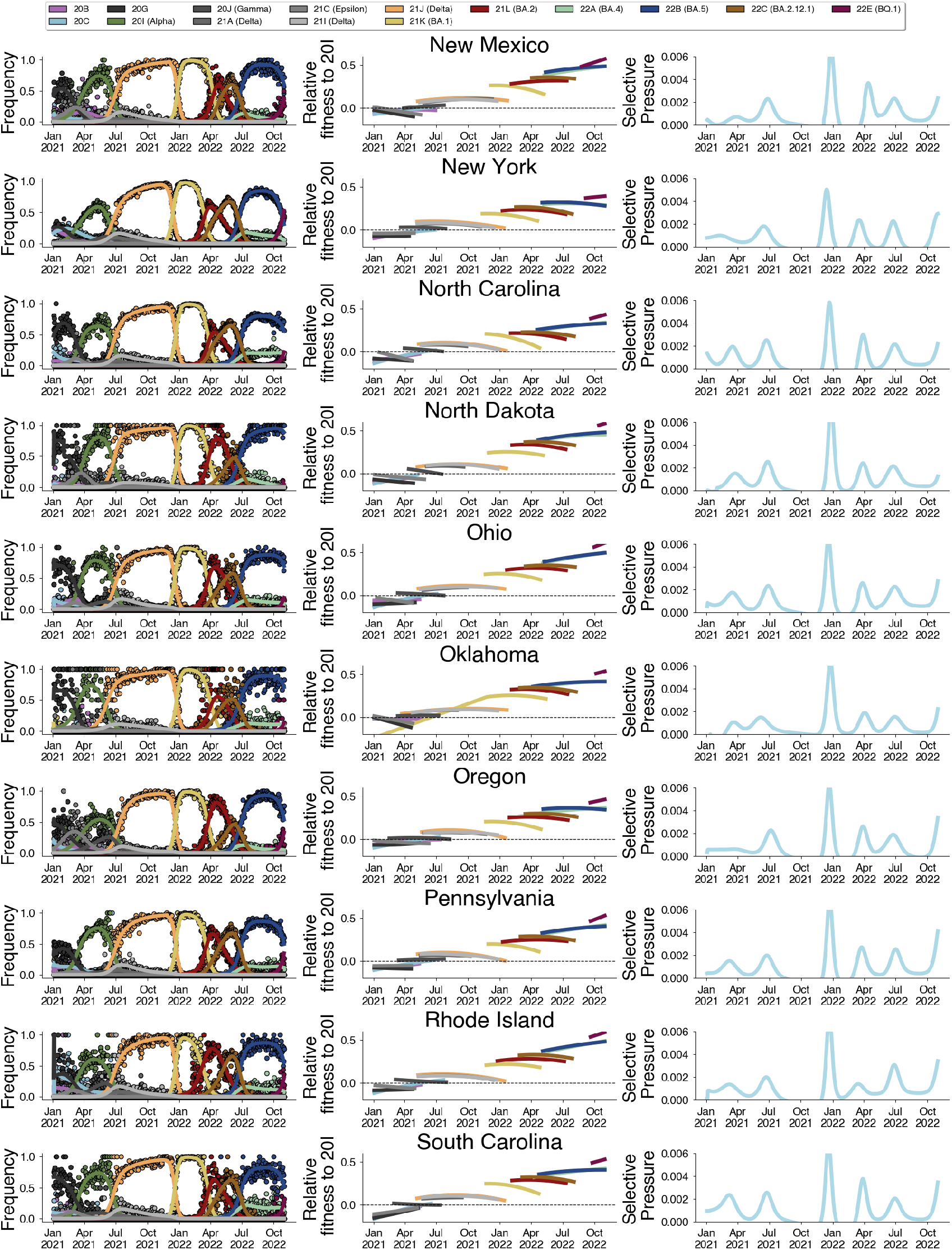
Estimated variant frequencies, relative fitnesses, and selective pressure. New Mexico to South Carolina.

**Fig. S8.**
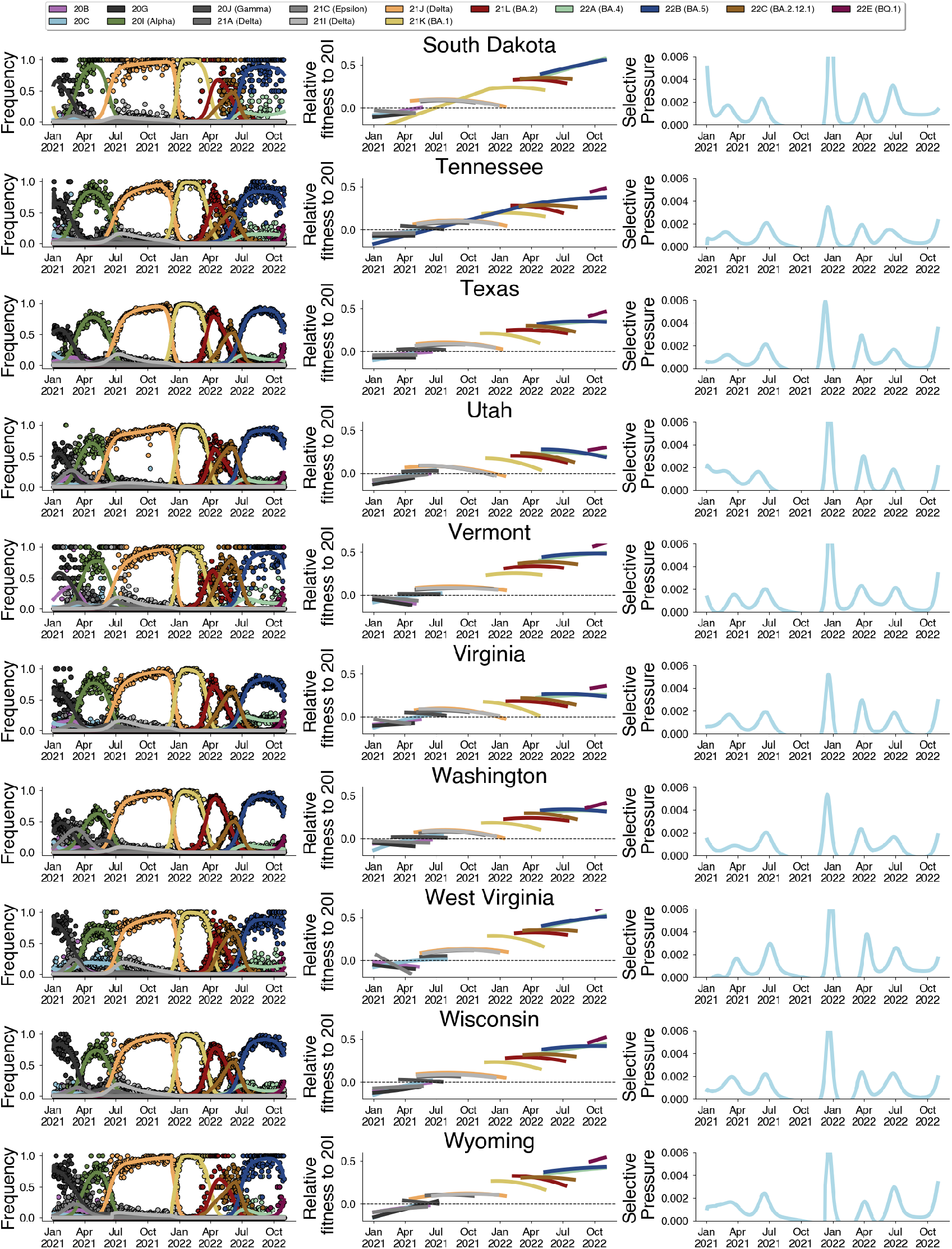
Estimated variant frequencies, relative fitnesses, and selective pressure. South Dakota to Wyoming.

**Fig. S9.**
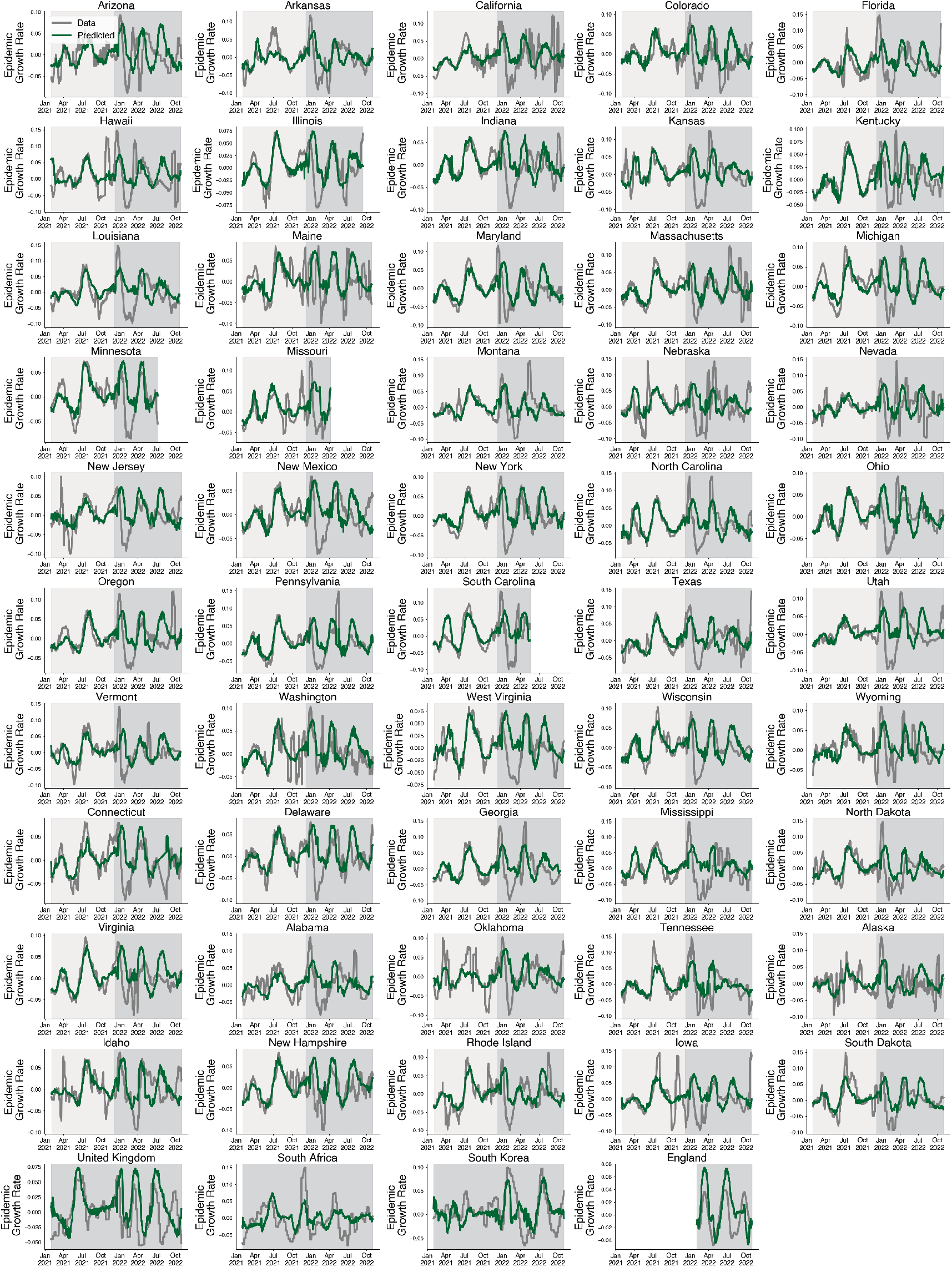
Predictions for empirical growth rate using selective pressure for all locations.

**Fig. S10.**
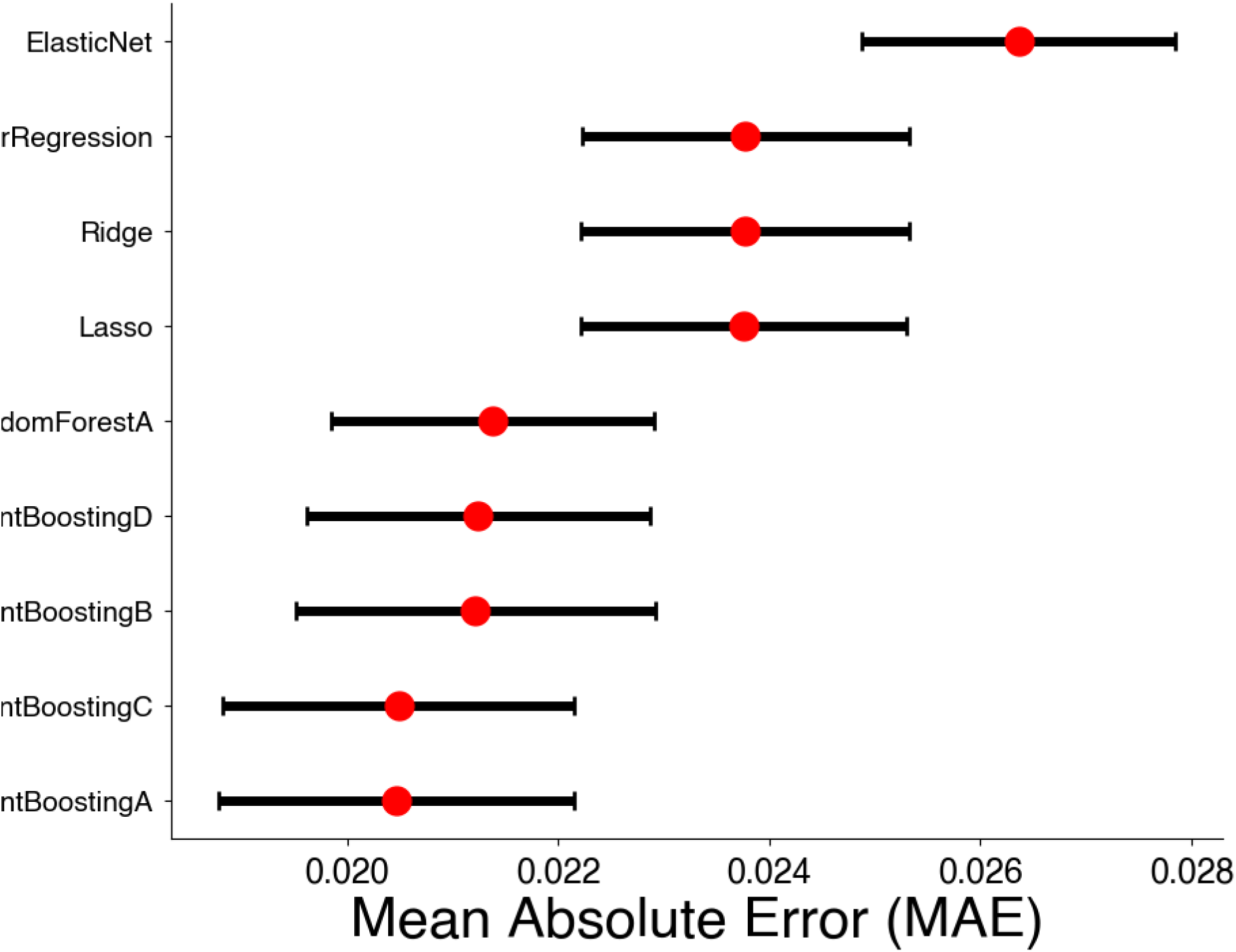
Cross-validation error by model. We compare the errors between models fit on 10 time series cross-validation splits.

**Fig. S11.**
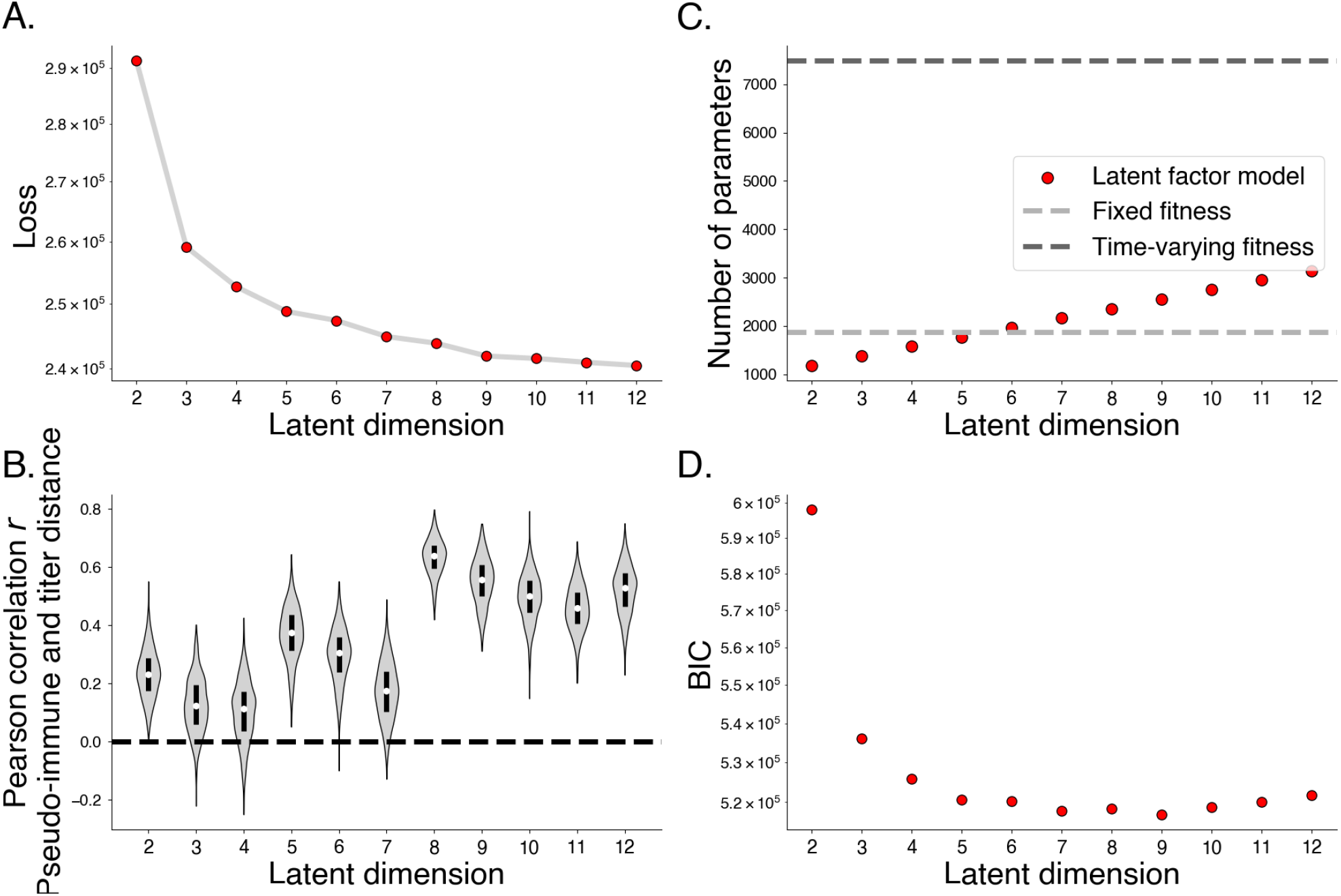
Comparing latent factor model by number of latent immune dimensions. A. Maximum a posteriori loss by number of latent immune dimensions. B. Spearman correlation between pseudo-immune and titer distance by number of latent immune dimensions. C. Number of parameters by number of latent immune dimensions. D. Bayesian Information Criterion (BIC) by number of latent immune dimensions.

**Fig. S12.**
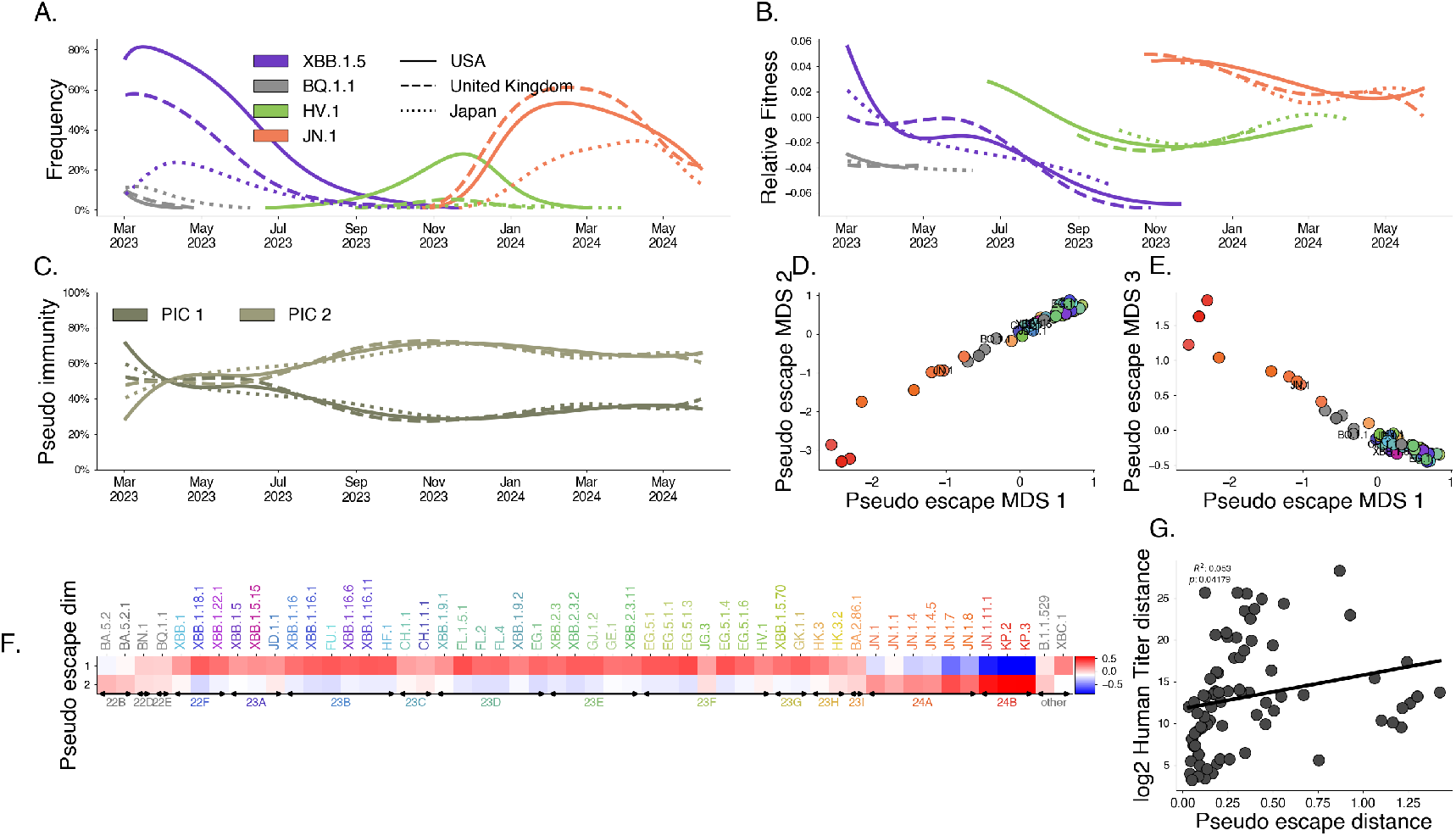
Latent factor model with *D* = 2 pseudo immune dimensions.

**Fig. S13.**
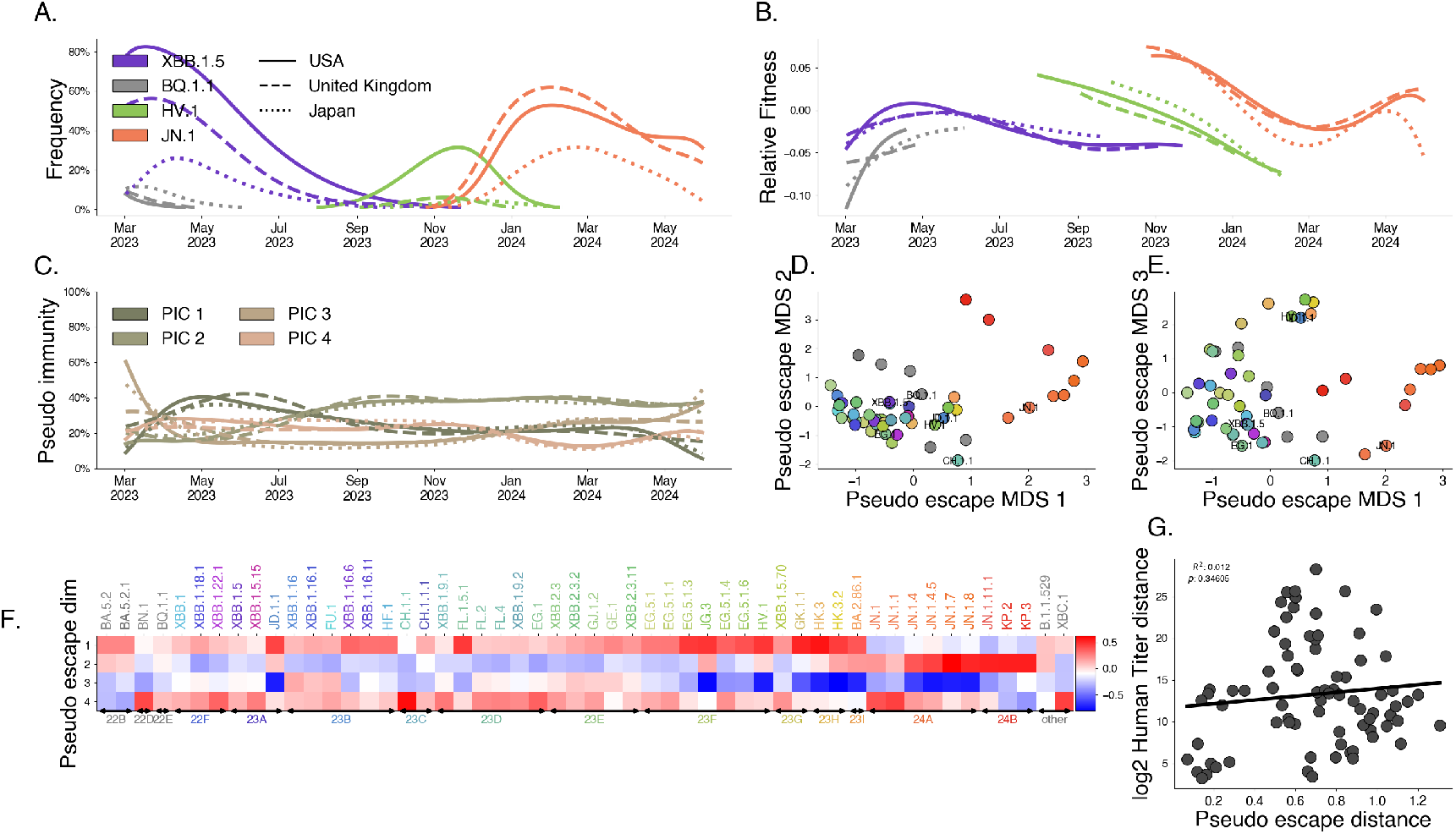
Latent factor model with *D* = 4 pseudo immune dimensions.

**Fig. S14.**
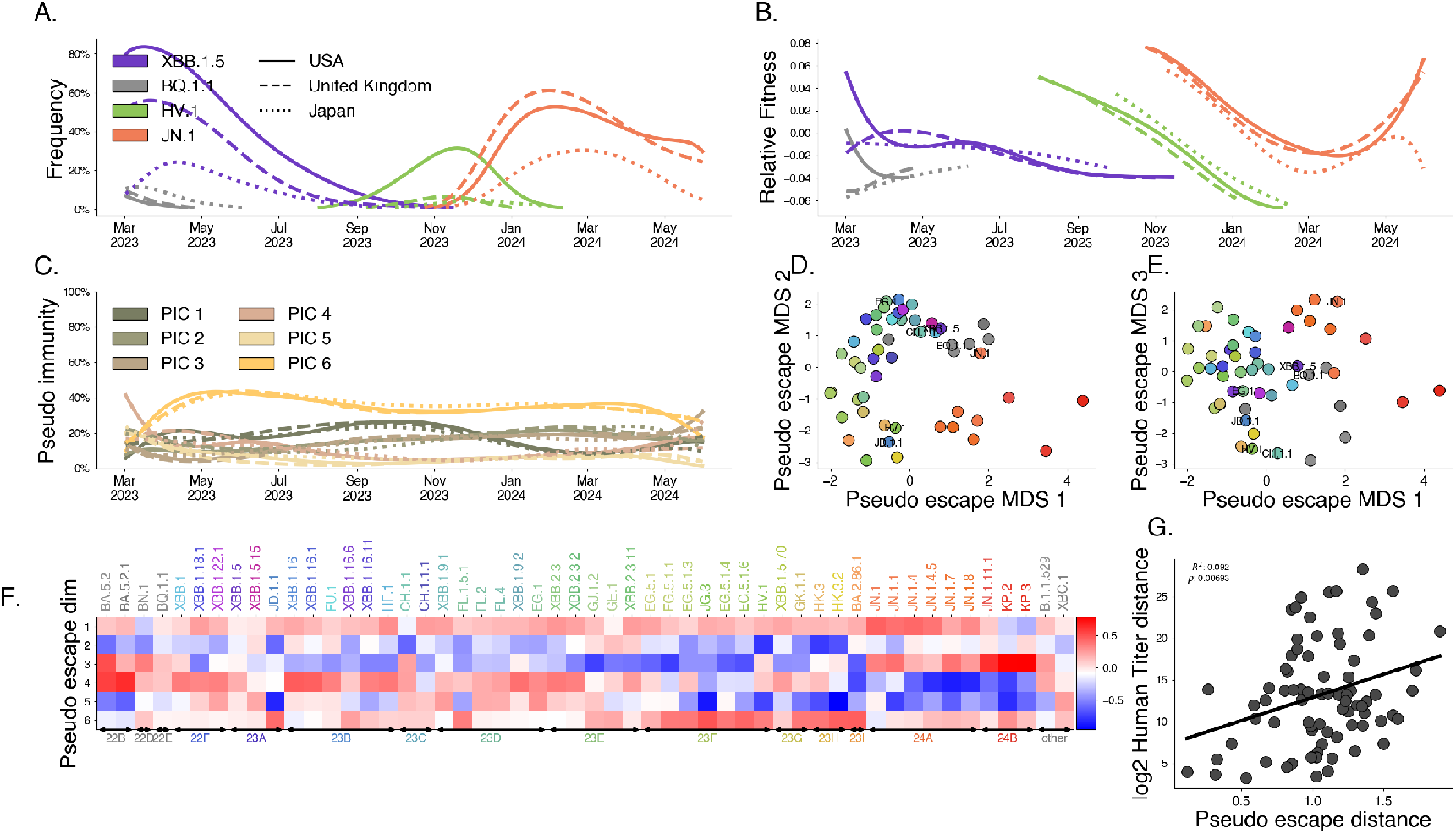
Latent factor model with *D* = 6 pseudo immune dimensions.

**Fig. S15.**
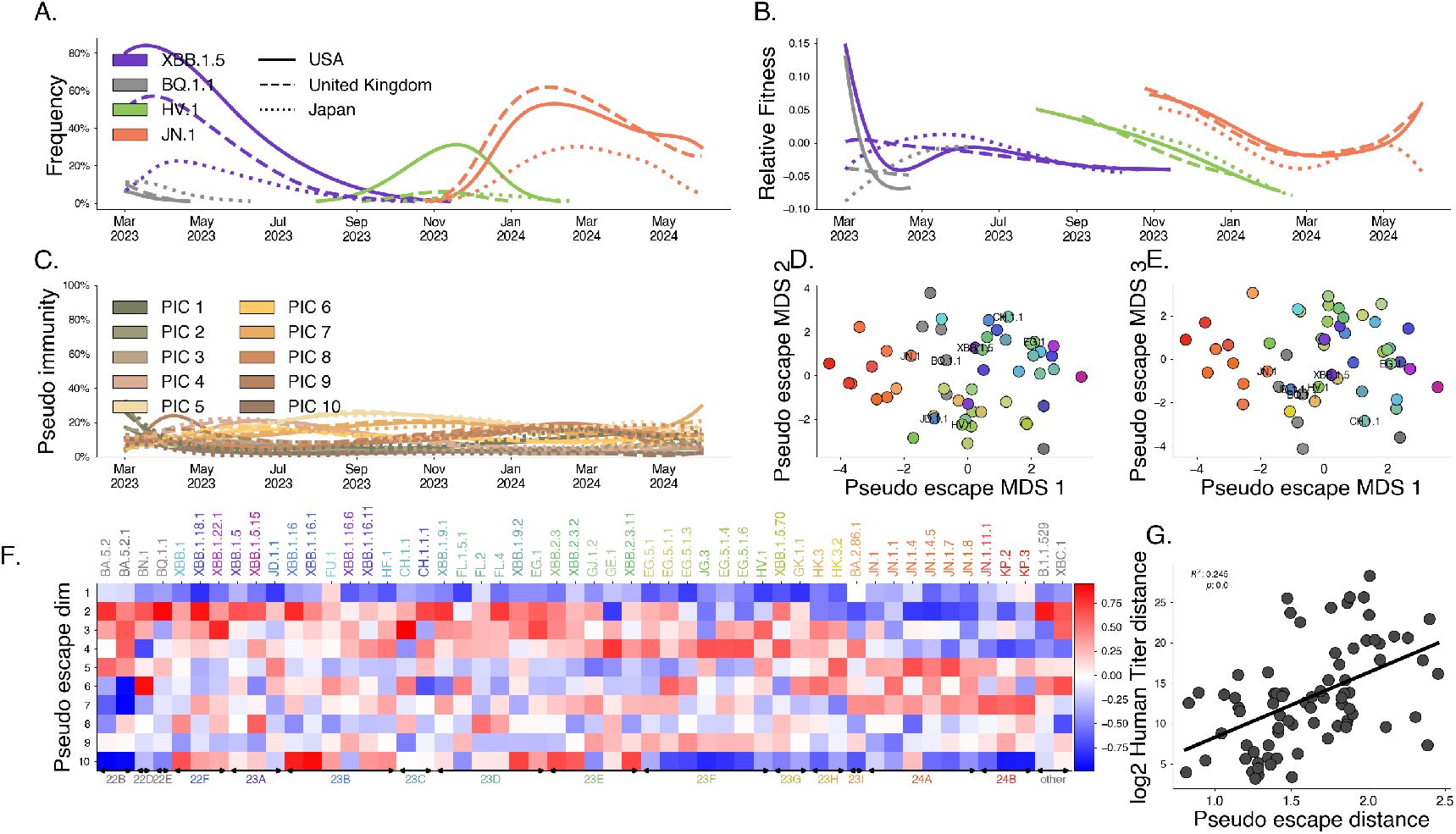
Latent factor model with *D* = 10 pseudo immune dimensions.

**Fig. S16.**
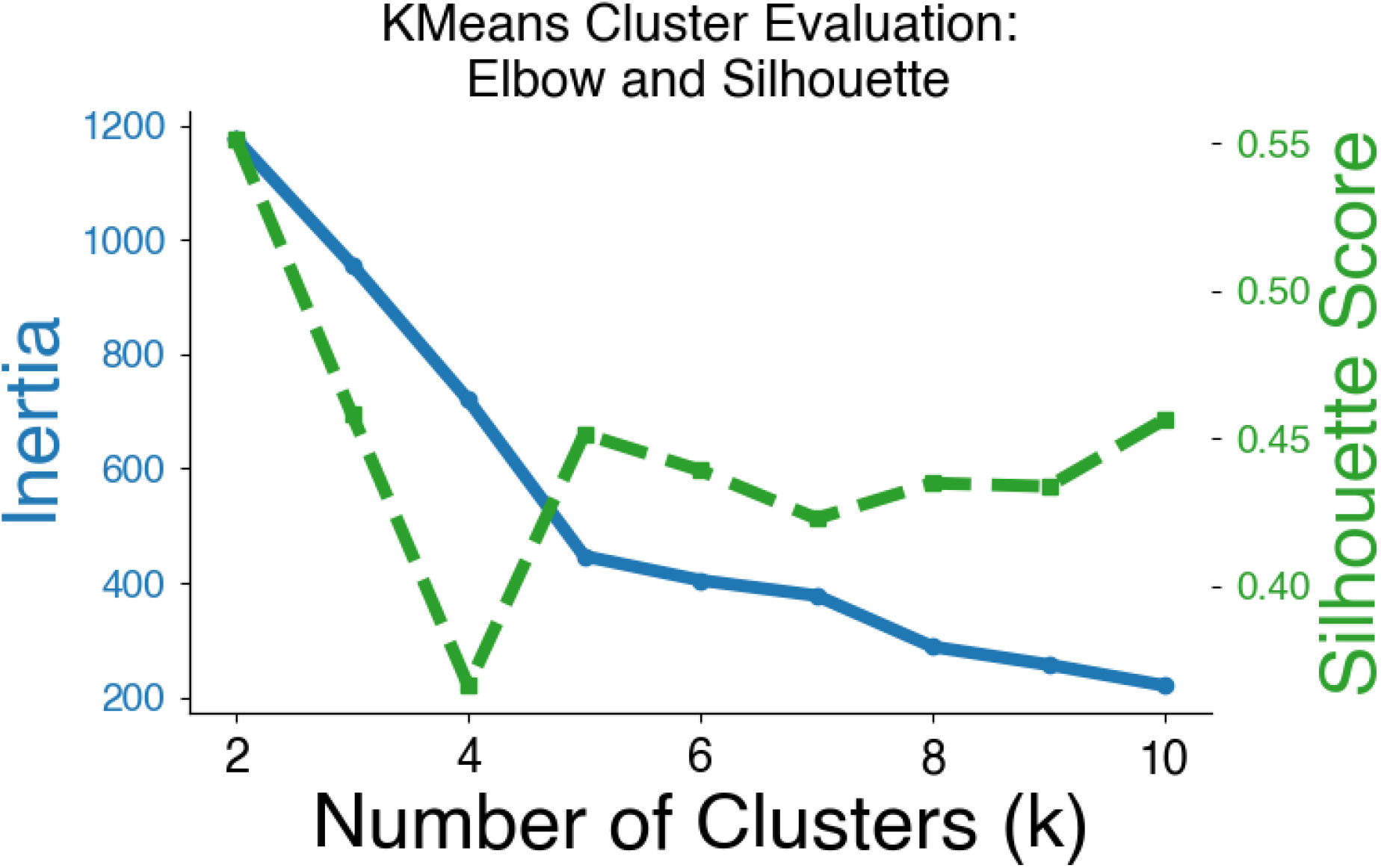
Choosing the number of immune clusters (*k* = 5) using inertia and silhouette score. This analysis uses only titer data from Jian et al. [7].

**Fig. S17.**
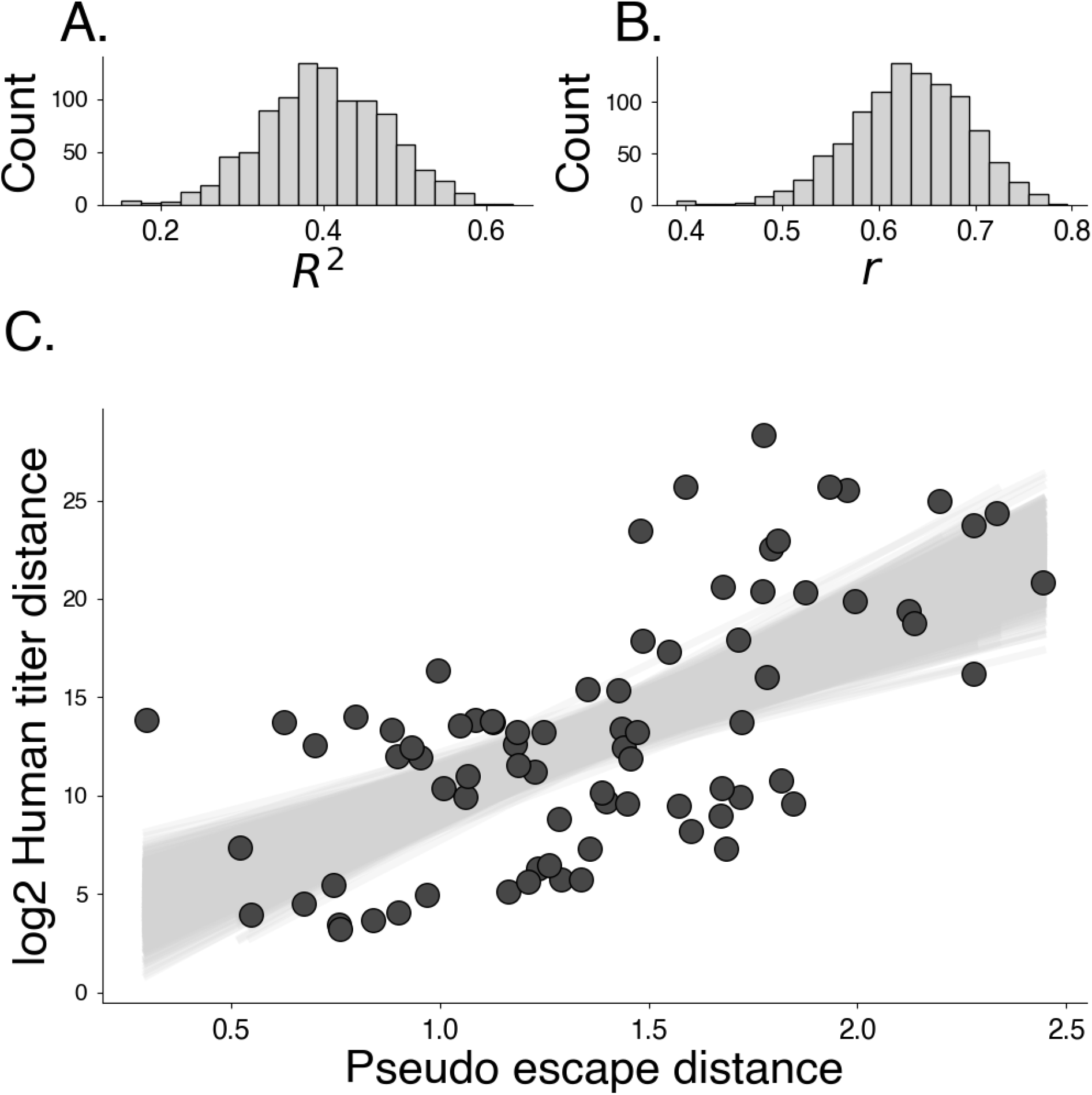
Bootstrapping pseudo-immune distance and human titer distance analysis (*N*_replicate_ = 1, 000).

**Fig. S18.**
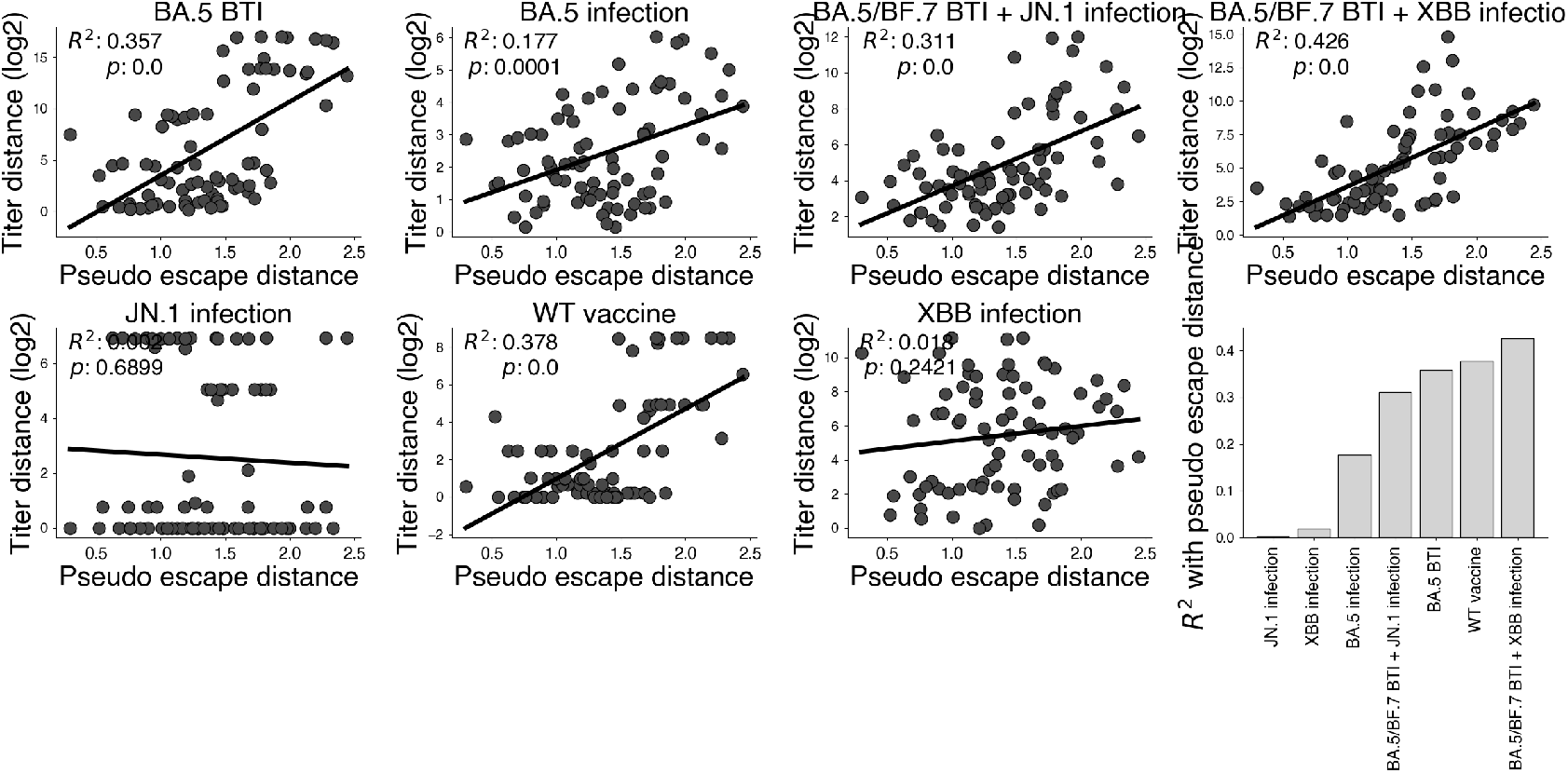
Comparing pseudo-escape distance and titer distance between exposure groups.

**Fig. S19.**
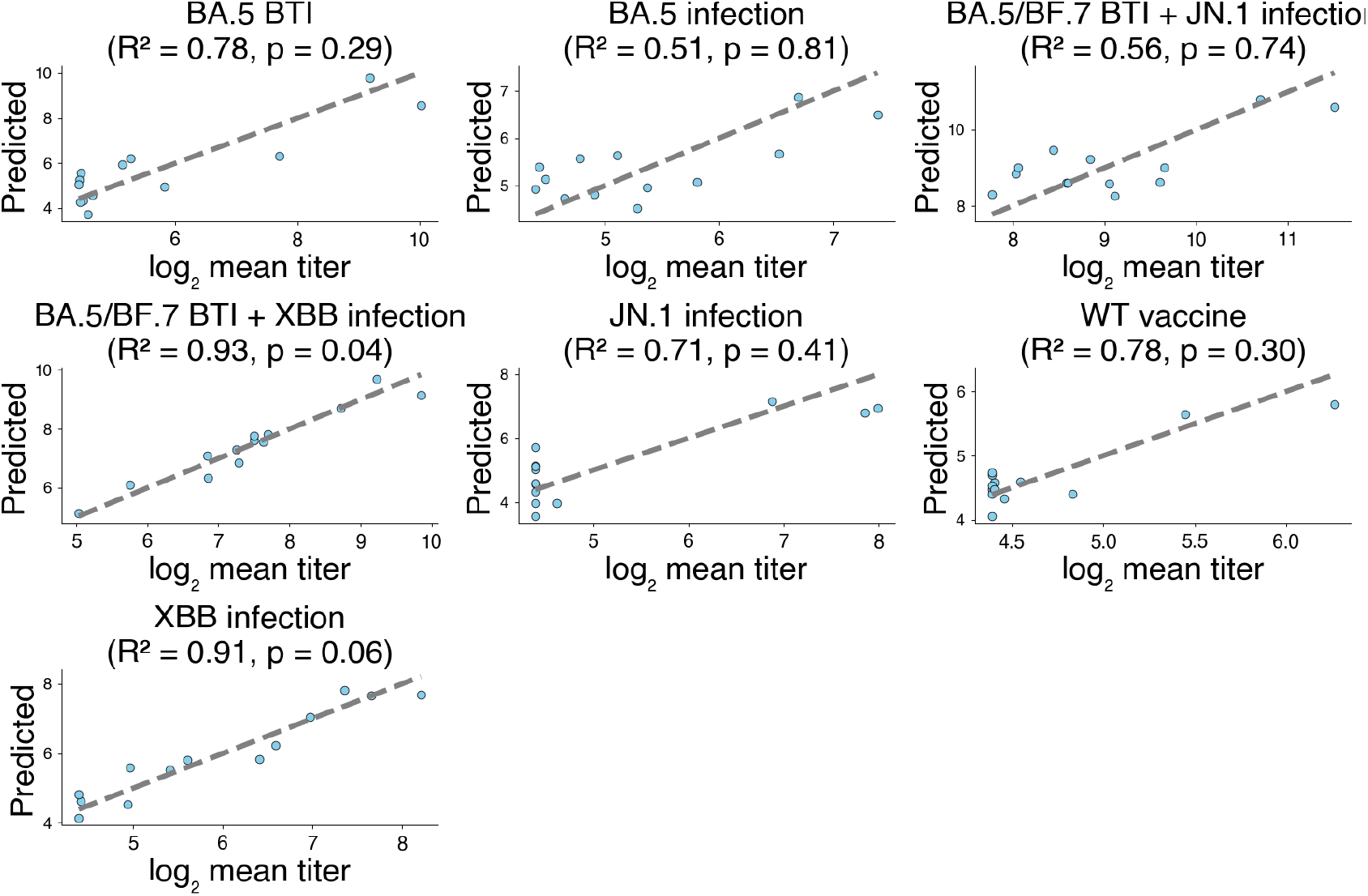
Titers predicted with pseudo-escape values and observed titers within infection cohorts.

**Fig. S20.**
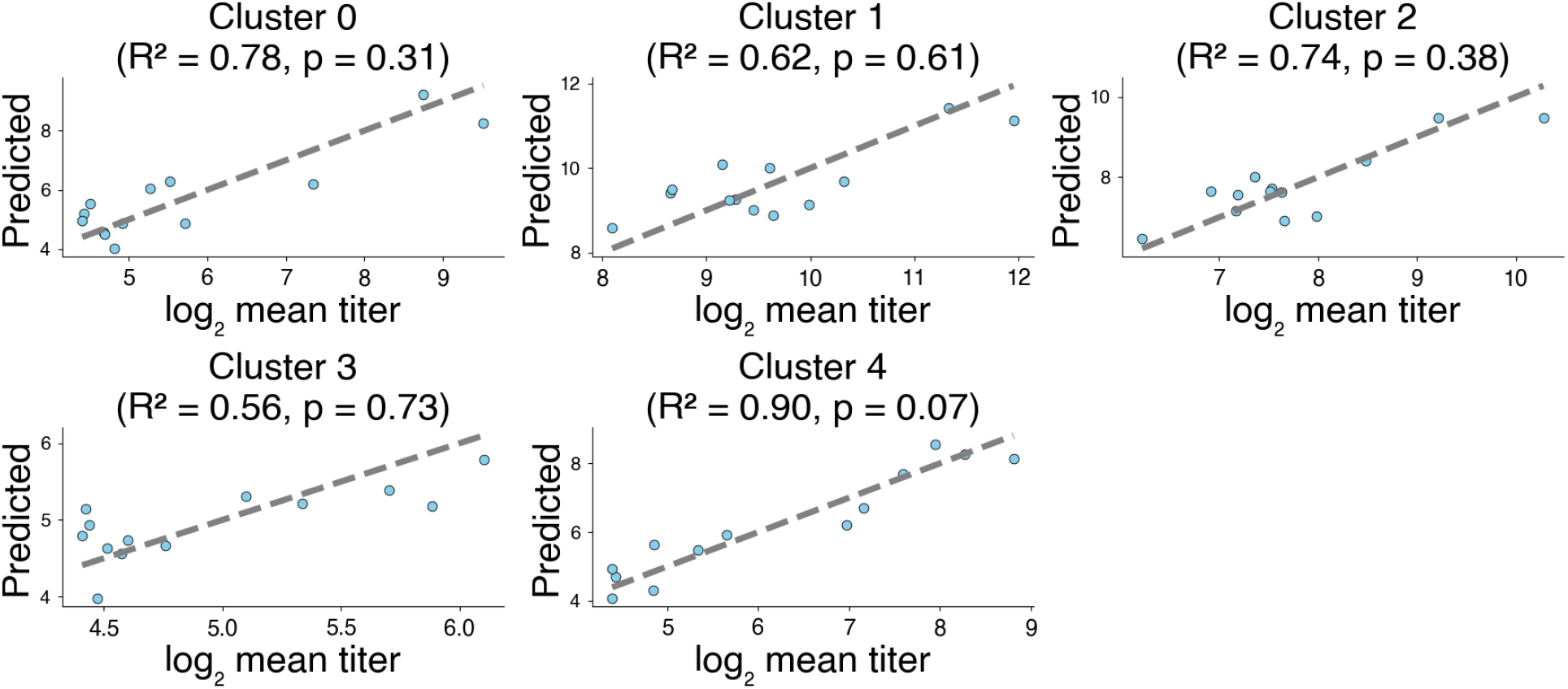
Titers predicted with pseudo-escape values and observed titers within immune clusters.

